# Defining normal inflammatory marker and vital sign responses to suspected bloodstream infection in adults with positive and negative blood cultures

**DOI:** 10.1101/2023.10.23.23297340

**Authors:** Qingze Gu, Jia Wei, Chang Ho Yoon, Kevin Yuan, Nicola Jones, Andrew Brent, Martin Llewelyn, Tim EA Peto, Koen B Pouwels, David W Eyre, A Sarah Walker

**Affiliations:** Nuffield Department of Medicine, University of Oxford, Oxford, UK; Oxford University Hospitals NHS Foundation Trust, Oxford, UK; Health Economics Research Centre, Nuffield Department of Population Health, University of Oxford, Oxford, UK; Big Data Institute, Nuffield Department of Population Health, University of Oxford, Oxford, UK; Brighton and Sussex Medical School, Brighton, UK; NIHR Health Protection Research Unit in Healthcare Associated Infections and Antimicrobial Resistance, University of Oxford, Oxford, UK

**Author notes:** Contribution considered equal (David W Eyre, A Sarah Walker).

## Abstract

**Background:** Patients respond differently to bloodstream infection (BSI) and associated antibiotic treatment, for many reasons, including different causative pathogens, sources of infection, and patient characteristics. This heterogeneity can hamper use of different clinical parameters to track treatment response as the same absolute values, or even change from presentation, may have different implications, depending on the expected trajectory, which is often incompletely understood.

**Methods:** We included patients ≥16y from Oxford University Hospitals (01-January-2016 to 28-June-2021) with any blood culture taken, grouping cultures into suspected BSI episodes (14-day de-duplication). We used linear and latent class mixed models to estimate trajectories in C-reactive protein (CRP), white blood count, heart rate, respiratory rate and temperature and identify subgroups with heterogenous CRP responses. Centile charts for expected CRP responses were constructed via the lambda-mu-sigma method.

**Findings:** 88,348 suspected BSI episodes occurred in 60,647 adults; 6,910(7.8%) were culture-positive with a probable pathogen (1,914[2.2%] Gram-positive, 3,736[4.2%] Gram-negative, 1,260[1.4%] other pathogens/polymicrobial), 4,307(4.9%) contained potential contaminants, and 77,131(87.3%) were culture-negative. Overall, CRP levels generally peaked between day 1-2 after blood culture collection, with varying responses for different pathogens and infection sources in adjusted models (interaction p<0.0001).

We identified five different CRP trajectory subgroups: peak on day 1 (36,091;46.3%) or 2 (4,529;5.8%), slow recovery (10,666;13.7%), peak on day 6 (743;1.0%), and low response (25,928;33.3%). 42,818(63.5%) culture-negative vs. 5,879(89.6%) pathogen-culture-positive episodes had acute response (day 1-2 peak/slow recovery). Centile reference charts constructed from those peaking on day 1-2 showed the same post-presentation CRP values and change from presentation reflected different responses depending on patients’ initial values.

**Interpretation:** Although infection sources and pathogens are associated with varying responses to BSI, there is distinct underlying heterogeneity in responses. The centile reference charts developed could facilitate more precise tracking of recovery, enable identification of patients not recovering as expected, and help personalise infection management.

**Research in context:** *Evidence before this study:* We searched PubMed up to 28 June 2023, for published English articles with the terms “response” AND (“pattern” OR “trend” OR “trajector*”) AND (“bloodstream infection” OR “sepsis”). No studies described pathogen-specific response trajectories for laboratory tests and vital signs. Several studies identified sepsis sub-phenotypes using group-based trajectory modelling based on trajectories of vital signs, white blood cell and Sequential Organ Failure Assessment score. Specifically, three studies identified four temperature trajectory subgroups using measurement within first 72h: “hyperthermic, slow resolvers”, “hyperthermic, fast resolvers”, “normothermic”, and “hypothermic”. One study identified seven different systolic blood pressure trajectory subgroups using measurements within 10h after hospitalisation and investigated their association with hospital mortality. One study identified seven white blood cell (WBC) count trajectories over the first seven days in the ICU and concluded rising trajectory was independently associated with increased mortality compared with the stable trajectory. Another study found four sub-phenotypes based on four different longitudinal vital signs from the first 8h of hospitalisation, including temperature, heart rate, respiratory rate, systolic and diastolic blood pressure. Several studies used Sequential Organ Failure Assessment score to identify trajectory subgroups, and they identified four or five subgroups using data from the first 72h or first 8 days. There were no published studies estimating expected C-reactive protein (CRP) response in standard responders.

*Added value of this study:* To our knowledge, this is the first study to characterise pathogen-specific and infection source-specific response trajectories of multiple clinical parameters, including CRP, WBC count, heart rate, respiratory rate, and temperature. We identified five different CRP trajectory subgroups and found that 42,818 (63.5%) of culture-negative vs. 5,879 (89.6%) of pathogen-culture-positive episodes had acute response, i.e. a peak in CRP on day 1 or 2 or a slow recovery, and that these CRP subgroups had equivalent parallel responses for the other clinical parameters. Centile reference charts (analogous to paediatric growth charts) were created based on the standard CRP responders (i.e., a peak in CRP on day 1 or 2, assuming that these reflected “normal” response to effective antibiotics). These can be used to standardise assessment of infection progression and treatment response in patients with suspected bloodstream infection given the heterogeneity in these responses. These reference charts could be useful to guide management independent of microbiological test results, e.g., prior to culture results becoming available.

*Implications of all the available evidence:* Patient characteristics and host responses are heterogeneous, both initially at presentation and throughout responses to infection, making it challenging to define a single “normal” response to culture-positive and culture-negative suspected bloodstream infection. By applying centile-based methods to large-scale electronic health records, we provide a visually intuitive means of assessing biomarker response, potentially aiding clinical decisions by allowing individual-level observations to be assessed against evidence-based references for expected recovery in patients treated with effective antibiotics, taking into account individual-level heterogeneity.

## Introduction

Effective treatment of bloodstream infections (BSI) and sepsis is an important priority.^1^ Management relies on timely initiation of active antimicrobials, but blood cultures identify a causative infectious agent in only 30–40% of serious infections.^1,2^ Hence, most antimicrobial treatment is started, and often continued, empirically. Such treatment may not always provide adequate coverage, with under-treatment leading to more severe infections and higher mortality;^3^ however wide use of empirical broad-spectrum antibiotics and escalation of empirical treatment in patients deemed not to be responding adequately significantly contributes to the growing global antimicrobial resistance crisis.^4^

Laboratory test results and clinical assessments can guide treatment decisions, especially when blood culture results remain unavailable/non-conclusive, with C-reactive protein (CRP),^5^ procalcitonin (PCT)^2,6^ and white blood cell (WBC) counts,^6^ and vital signs including temperature, heart rate and respiratory rate,^7^ routinely monitored to assess status. Scoring systems, including the Systemic Inflammatory Response Syndrome (SIRS) criteria, National Early Warning Score (NEWS), and Sequential Organ Failure Assessment (SOFA) Score, can provide key insights into status and risk of deterioration.^1^ However, patient characteristics and host responses are heterogeneous, both initially at presentation and throughout infections,^7,8^ making it challenging to determine whether an individual patient’s response to treatment of a suspected BSI is “normal”.

Detailed electronic health records (EHRs), combined with advanced statistical approaches such as latent class mixed models (LCMM),^9^ potentially allow identification of different patient response trajectories and underlying heterogeneity. Additionally, centile-based methods, as used in paediatric growth charts,^10^ could be used to construct reference expected clinical responses given a patient’s status at presentation and effective treatment. These could be used to identify deviations from a typical recovery trajectory to inform individualised clinical decision-making. Previous studies used group-based models to identify subgroups of patients with different vital signs, WBC and SOFA score trajectories in patients with suspected sepsis,^7,11–18^ however, to date, none have applied centile-based methods to infection responses.

We therefore aimed: first, to estimate changes in routinely collected clinical parameters following negative or positive blood cultures, stratified by pathogen/clinical syndrome; second, to identify underlying response patterns using latent class trajectory modelling, to identify those responding standardly to (effective) antibiotics; and third, to construct centile reference charts for expected clinical response in standard responders to support clinicians tailoring treatment to individual patient responses.

## Methods

We used de-identified data from the Infections in Oxfordshire Research Database (IORD), containing information from all inpatient admissions at the Oxford University Hospitals NHS Foundation Trust (OUH), United Kingdom, together with vital signs, microbiology and biochemistry/haematology results and antibiotics prescribed in hospital. OUH contains ∼1000 beds in four hospitals, providing all acute care and pathology services to a population of ∼750,000 and specialist services to the surrounding region. Ethical approval was obtained from the National Research Ethics Service South Central Oxford C Research Ethics Committee (19/SC/0403) and the national Confidentiality Advisory Group (19/CAG/0144).

Patients ≥16y and with ≥1 blood culture taken were included. A new suspected infection episode was defined if there were >14 days since the last blood culture, prioritising pathogens, then contaminants, then any negative cultures as the index blood culture to define the start of each episode (date/time of the blood collection for culture). Episodes with index blood cultures taken >24h before admission or after discharge were excluded.

### Statistical analyses

Linear mixed models were used to estimate CRP, WBC and vital signs (heart rate, respiratory rate, tympanic temperature) trajectories throughout suspected BSI episodes from −1 day (CRP, WBC) or −6 hours (vital signs) before to +8 days after the start of each episode, using natural cubic splines to allow for non-linearity over time, adjusting for presumed source of infection (identified from antibiotic prescribing indications^19^), community-onset (episode start ≤48 hours after admission), blood culture result (positive, potential contaminant, negative) and pathogen group (based on genus and clinical significance, **Table S1**), age, sex, Charlson and Elixhauser scores and immunosuppression, and their interaction with time if interaction-p<0.05.

Separate adjusted models were fitted to examine effects of source of infection and baseline antimicrobial susceptibility on response trajectories (determined by laboratory tests and information on intrinsic resistance^20^).

We did not adjust for updates to antibiotics over time following baseline because of potential time-dependent confounding. Instead, we used unadjusted latent class mixed models (LCMM) to identify underlying population-level heterogeneity in CRP response trajectories (**see supplement**) and hence identify those responding standardly to (effective) antibiotics by assigning each episode to the class with the highest posterior probability. This approach has the advantage that culture-negative episodes can also be considered. Hospital empirical antibiotic recommendations are based on susceptibilities data from recent previous infections, with antibiotic treatment switched promptly if a resistant pathogen is identified. However, many infections are culture negative, such that resistant infections may be missed, and furthermore it may take several days to identify culture-positive resistant infections.

Centile reference charts for expected CRP response in standard responders with peak response on day 1-2 were constructed using the lambda-mu-sigma (LMS) method^10^ (**see supplement**).

## Results

From 1-January-2016 to 28-June-2021, 24.4% (95,928/392,443) of admissions had blood cultures taken during their hospital stay (39.5% [82,535/208,699] of emergency and 4.4% [7,132/163,201] of elective admissions; overall 122 blood cultures per 1,000 patient-days). There were 88,348 suspected BSI episodes in 60,647 patients (**Figure S1**); a single Gram-positive pathogen was identified in 1,914 (2.2%), a single Gram-negative pathogen in 3,736 (4.2%), 1,260 (1.4%) had other pathogens or were polymicrobial, 4,307 (4.9%) had only a potential contaminant, and 77,131 (87.3%) were culture-negative (**Table 1**). At the start of each episode, the median age was 67.3 (IQR 48.5–80.4) years. Patients had relatively few comorbidities (median Charlson 1 (IQR 0–2)), with only 12802 (14.5%) episodes in immunosuppressed patients; most episodes were community-onset (71,258, 80.7%). Classifying source from clinician-recorded antibiotic prescribing indications, most episodes had respiratory (22,818; 25.8%), multiple (11,012; 12.5%), urinary (9,275; 10.5%), abdominal (6,912; 7.8%) and skin, soft tissue and orthopaedic (6,297; 7.1%) sources (non-specific sources in 27,964[31.7%] episodes).

**Table 1.** Characteristics at the start of 88,348 suspected bloodstream infection (BSI) episodes between 01-January-2016 and 28-June-2021. Percentages in the header are of all episodes, and in the main body are column percentages within each group; continuous variables are summarised using median (IQR). Baseline NEWS score were calculated using the closest set of vital signs within 1 day before to 1 day after the start of each episode.

| Characteristic | Gram-positive Pathogens,<br>N = 1 914 (2.2%)<sup>1</sup> | Gram-negative Pathogens,<br>N = 3 736 (4.2%)<sup>1</sup> | Other Pathogens,<br>N = 1 260 (1.4%)<sup>1</sup> | Potential Contaminant(s),<br>N = 4 307 (4.9%)<sup>1</sup> | Culture-negative,<br>N = 77 131 (87.3%)<sup>1</sup> | Overall,<br>N = 88 348 (100%)<sup>1</sup> |
| --- | --- | --- | --- | --- | --- | --- |
| Age at admission (years) | 70·6 (54·4, 82·1) | 74·9 (61·7, 84·1) | 65·3 (48·5, 78·9) | 69·0 (52·8, 81·2) | 66·6 (47·4, 80·1) | 67·3 (48·5, 80·4) |
| Sex (male) | 1 □ 146 (59·9%) | 2 □ 080 (55·7%) | 691 (54·8%) | 2 □ 213 (51·4%) | 37 □ 558 (48·7%) | 43 □ 688 (49·4%) |
| Ethnicity |  |  |  |  |  |  |
| White | 1 □ 591 (83·1%) | 3 □ 044 (81·5%) | 980 (77·8%) | 3 □ 447 (80·0%) | 61 □ 647 (79·9%) | 70 □ 709 (80·0%) |
| Other | 64 (3·3%) | 178 (4·8%) | 70 (5·6%) | 261 (6·1%) | 4 □ 631 (6·0%) | 5 □ 204 (5·9%) |
| Unknown | 259 (13·5%) | 514 (13·8%) | 210 (16·7%) | 599 (13·9%) | 10 □ 853 (14·1%) | 12 □ 435 (14·1%) |
| Charlson score | 1 (1, 2) | 2 (1, 3) | 1 (0, 2) | 1 (0, 3) | 1 (0, 2) | 1 (0, 2) |
| Elixhauser score | 3 (2, 5) | 3 (2, 5) | 3 (1, 4) | 3 (1, 4) | 2 (1, 4) | 2 (1, 4) |
| NEWS score (baseline) | 4 (2, 6) | 4 (2, 6) | 3 (2, 6) | 3 (1, 5) | 2 (1, 4) | 3 (1, 5) |
| Unknown | 131 (6·8%) | 210 (5·6%) | 117 (9·3%) | 442 (10·3%) | 6 □ 684 (8·7%) | 7 □ 584 (8·6%) |
| Immunosuppression | 311 (16·2%) | 719 (19·2%) | 306 (24·3%) | 661 (15·3%) | 10 □ 805 (14·0%) | 12 □ 802 (14·5%) |
| Diabetes mellitus | 490 (25·6%) | 972 (26·0%) | 254 (20·2%) | 971 (22·5%) | 14 □ 415 (18·7%) | 17 □ 102 (19·4%) |
| Palliative care | 179 (9·4%) | 359 (9·6%) | 144 (11·4%) | 308 (7·2%) | 3 □ 986 (5·2%) | 4 □ 976 (5·6%) |
| Community-onset | 1 □ 507 (78·7%) | 2 □ 852 (76·3%) | 863 (68·5%) | 3 □ 072 (71·3%) | 62 □ 964 (81·6%) | 71 □ 258 (80·7%) |
| Source of infection |  |  |  |  |  |  |
| Respiratory | 432 (22·6%) | 378 (10·1%) | 185 (14·7%) | 1 □ 179 (27·4%) | 20 □ 644 (26·8%) | 22 □ 818 (25·8%) |
| Multiple sources | 422 (22·0%) | 817 (21·9%) | 228 (18·1%) | 565 (13·1%) | 8 □ 980 (11·6%) | 11 □ 012 (12·5%) |
| Urinary | 132 (6·9%) | 1 □ 044 (27·9%) | 116 (9·2%) | 427 (9·9%) | 7 □ 556 (9·8%) | 9 □ 275 (10·5%) |
| Abdominal | 101 (5·3%) | 590 (15·8%) | 169 (13·4%) | 254 (5·9%) | 5 □ 798 (7·5%) | 6 □ 912 (7·8%) |
| Skin, soft tissue,<br>orthopedic | 272 (14·2%) | 88 (2·4%) | 80 (6·3%) | 261 (6·1%) | 5 □ 596 (7·3%) | 6 □ 297 (7·1%) |
| CNS | 35 (1·8%) | 23 (0·6%) | 21 (1·7%) | 72 (1·7%) | 1 □ 063 (1·4%) | 1 □ 214 (1·4%) |
| Other | 117 (6·1%) | 60 (1·6%) | 87 (6·9%) | 160 (3·7%) | 2 □ 432 (3·2%) | 2 □ 856 (3·2%) |
| Unspecific | 403 (21·1%) | 736 (19·7%) | 374 (29·7%) | 1 □ 389 (32·2%) | 25 □ 062 (32·5%) | 27 □ 964 (31·7%) |

<sup>1</sup>Median (IQR); n (%)

### CRP response trajectories following negative/positive blood cultures

77,957 (88.2%) suspected BSI episodes in 54,381 (89.7%) patients had ≥1 CRP measurement available (median 4 (IQR 2–6, range 1–20) measurements per episode). The distribution of blood culture results/pathogen groups was broadly similar between episodes with and without CRP measurements (standardised mean difference (SMD) <0.22, **Table S1**) except slightly fewer culture-negative results in those with CRP results (SMD=0.22). CRP levels increased sharply and generally peaked between day 1 and day 2 post blood-culture collection, with varying rates of increase and peaks for different pathogen groups (interaction p<0.0001, **Figure 1**). Adjusted CRP response trajectories differed most substantially in Gram-positive infections (**Figure 1A**), with *Streptococcus pneumoniae* infections rising much faster than other Gram-positive (or Gram-negative) pathogens and peaking at day 1 (∼290mg/L), followed by rapid declines and near stability by day 6 (∼65mg/L). CRP also increased rapidly with beta-haemolytic Streptococci but peaked slightly later, reaching ∼240mg/L on day 1.3 and then decreasing rapidly (to ∼50mg/L by day 8). CRP response trajectories for Gram-negative infections were broadly similar, peaking at 175–215mg/L after day 1 before falling back to ∼35mg/L (**Figure 1B**). For other pathogens, peak CRP levels were higher in episodes with anaerobic and polymicrobial infections (190–200mg/L), and the latter had the slowest recovery rate, remaining at ∼75mg/L by day 8; recovery was also slower in *Candida* episodes (∼60 mg/L by day 8, **Figure 1C**). CRP responses were still seen in those with only potential contaminants or no organism identified, and were similar, CRP peaked at 95–115mg/L after just over day 1 (**Figure 1D**).

**Figure 1.**
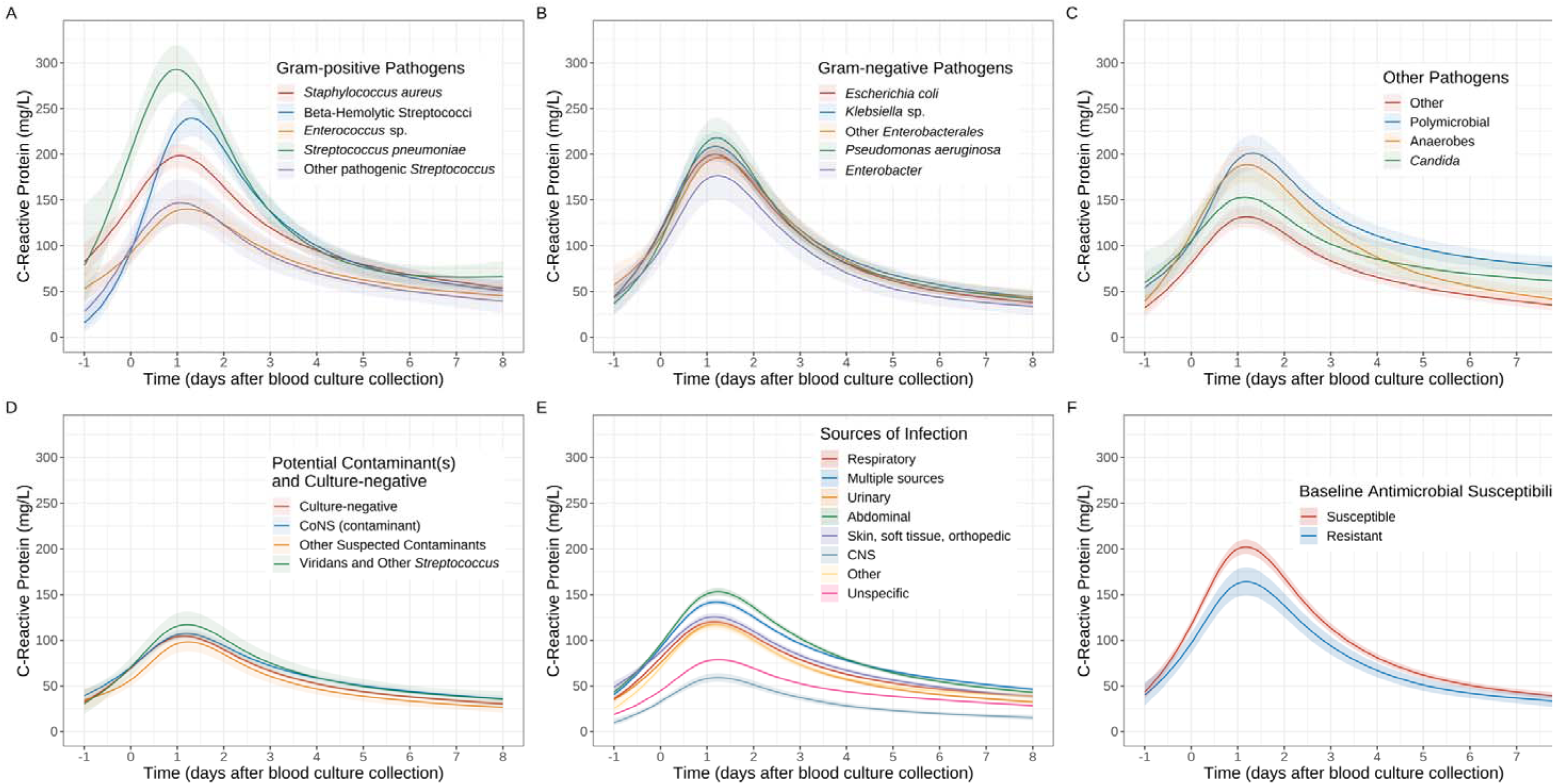
CRP response trajectories following different blood culture results (Gram-positive pathogens (A), Gram-negative pathogens (B), other pathogens (C), and potential contaminants and culture-negative results (D); adjusted for source of infection and other covariates), sources of infection (E) (not adjusted for blood culture results but adjusted for other covariates) and baseline antimicrobial susceptibilities (F) (adjusted for blood culture results, source of infection and other covariates). See **Figure S2A** for response trajectories of no baseline antimicrobial recorded and unknown baseline susceptibility. Predictions are plotted at the reference values of other adjusting variables: age = 64 years, male, Charlson score = 1, Elixhauser score = 3, community-onset, absence of immunosuppression, urinary source (excluding panel E), and *E. coli* infection (panel F only). Nonlinear trends were incorporated via natural cubic splines with four knots at the 20th, 40th, 60th and 80th percentiles of observed time values (day 0, day 0.8, day 2.4, day 4.7).

We considered associations with sources of infection in separate adjusted models not including pathogen/organism group (since this is not known for all patients whereas clinical syndromes largely are). The magnitude of differences between most sources was smaller than between pathogen groups (**Figure 1E**). Episodes with abdominal or multiple source(s) elicited the strongest CRP responses, with levels reaching ∼150mg/L and ∼130mg/L respectively by day 1. CRP responses were relatively weak for episodes with neurological and non-specific origin, with peaks of ∼60mg/L and ∼80mg/L. Episodes with the remaining origins had similar CRP responses to each other, peaking at 115–125mg/L. We also investigated associations with baseline antimicrobial susceptibilities in culture-positive infections in a model adjusted for pathogen groups and sources of infection. Episodes with pathogens susceptible to baseline antimicrobials elicited higher CRP responses than in those resistant to initial treatment (∼200mg/L vs. ∼165mg/L on day 1.2, **Figure 1F**; **Figure S2**; **Table S2**)

Additionally, after adjusting for infection source and pathogen, CRP levels were higher in males (∼20mg/L higher peak versus females, **Figure S3A**) and in episodes with nosocomial onset (20–60mg/L higher over the episode versus community-onset, time-interaction p<0.0001, **Figure S3B**). Compared to episodes in non-immunosuppressed patients, peak CRP levels were slightly lower in episodes in immunosuppressed patients and decreased more slowly over time (time-interaction p<0.0001, **Figure S3C**). CRP levels were also higher in older patients up to 70 years (∼9mg/L higher per 10 years older, **Figure S3D**). CRP peak levels were also slightly lower in episodes with higher Charlson comorbidity scores (p<0.001, **Figure S3E**).

### Response trajectories for other physiological measurements

Similar adjusted associations between blood culture results and response trajectories during suspected BSI episodes were observed for other physiological measurements, although to a lesser extent than for CRP (**Figures S4–7**). WBC peaked earlier than CRP, whereas heart rate, respiratory rate and temperature all declined rapidly over the first day following the start of the episode: however, differences associated with different pathogen groups were consistent. Episodes with *S. pneumoniae* and beta-haemolytic Streptococci had the highest initial heart rate, respiratory rate and temperature, at ∼105 beats/minute, 22–23 breaths/minute and 37.9–38.2°C 6h before blood culture collection, dropping to ∼83 beats/minute, ∼18 breaths/minute and ∼36.7°C by day 2 (**Figures S4A/S5A/S6A**); and the highest WBC count, peaking around the time of blood culture collection at ∼16×10^9^/L (**Figure S7A**). Similar to CRP, recovery was slower in patients with *Candida* and polymicrobial infections (**Figures S4C/S5C/S6C/S7C**), but response trajectories for other pathogen groups and sources of infection (**Figures S4E/S5E/S6E/S7E**) were broadly similar to each other. There was little difference in response trajectories for other physiological measurements between susceptible and resistant baseline treatments compared with CRP (**Figure S4F/S5F/S6F/S7F**).

### Latent classes of CRP response trajectories

From the 77,957 suspected BSI episodes with any CRP measurements, latent class modelling identified five different underlying subgroups of CRP response (**Figure 2A**, **Table 2**, **Figure S8**). These were distinguished by having their peak on day 1 (36,091[46.3%]), peak on day 2 (4,529[5.8%]), slow recovery (10,666[13.7%]), peak on day 6 (743[1.0%]) and low values throughout (25,928[33.3%]). Overall, 42,818 (63.5%) of culture-negative episodes and 2,589 (65.9%) episodes only with potential contaminants still had an acute CRP response (CRP peaking on day 1/2 or slow recovery) vs. 5,879 (89.6%) episodes with any pathogen (**Figure 2B**). For culture-pathogen-positive episodes with susceptibility results, 67.7% (3,580/5,286) with susceptible baseline antimicrobials had peak CRP on day 1/2 followed by typical recovery, vs. 55.7% (330/592) with resistant baseline treatment (**Figure 2C**).

**Figure 2.**
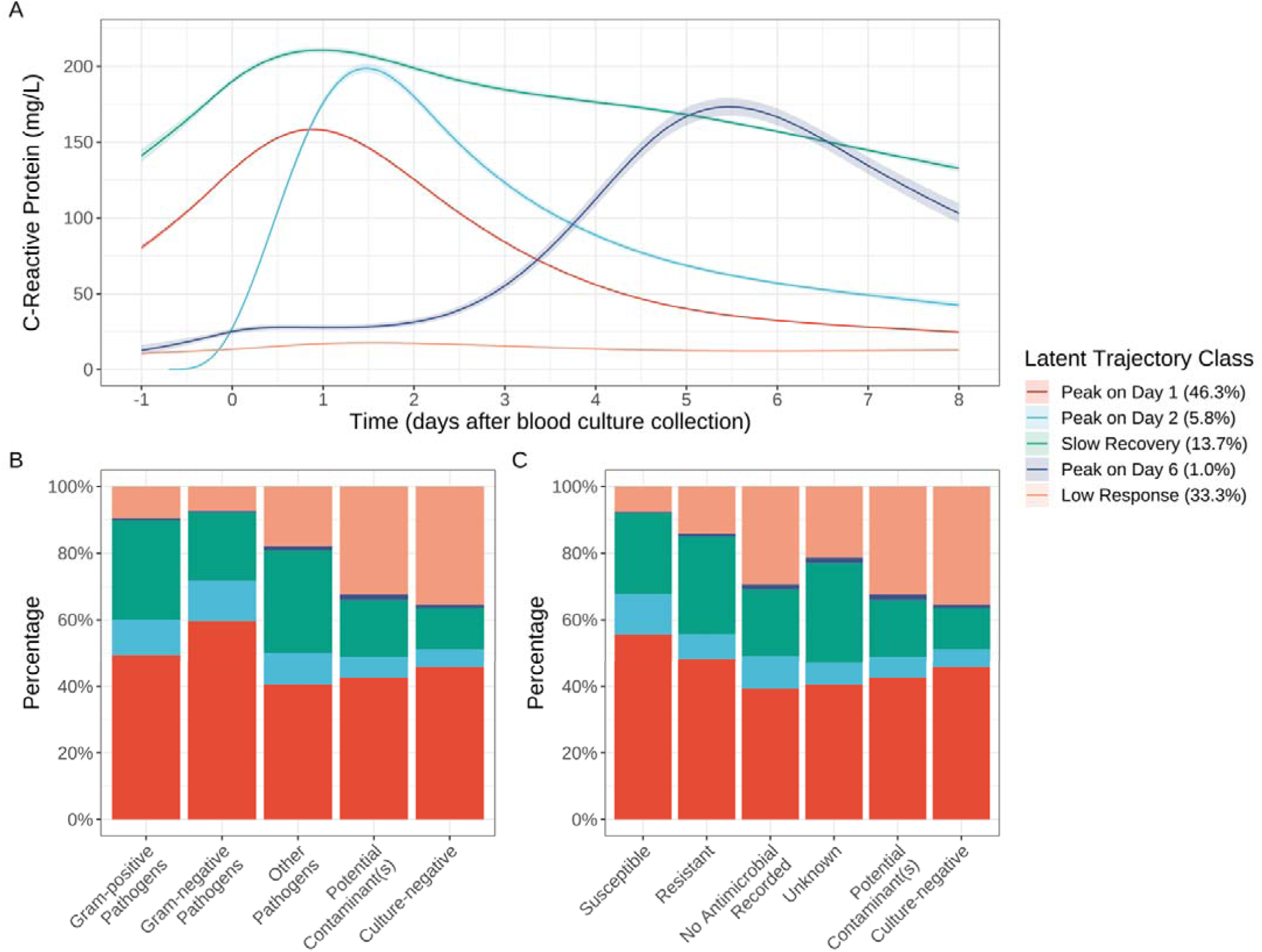
Latent classes of CRP response trajectories (A) (unadjusted for other covariates), distribution of the latent trajectory classes by pathogens identified (B) and baseline antimicrobial susceptibility (C). See **Figures S9B** and **S9D** for the distribution of blood culture results and baseline antimicrobial susceptibilities across each latent trajectory group.

**Table 2.** Characteristics of 77,957 suspected BSI episodes (with ≥1 measurement of CRP within 1 day before to 8 days after the start of each episode) by predicted latent trajectory class. See **Table S1** for comparison of pathogens isolated from included vs excluded episodes. The percentages in the header are of all episodes included, and in the main body are column percentages out of the total number of episodes within each distinct latent class; continuous variables are summarised using median (IQR). See supplement for definition of baseline antimicrobial susceptibility.

| Characteristic | Peak on Day 1,<br>N = 36 091 (46.3%)<sup>1</sup> | Peak on Day 2,<br>N = 4 529 (5.8%)<sup>1</sup> | Slow Recovery,<br>N = 10 666 (13.7%)<sup>1</sup> | Peak on Day 6,<br>N = 743 (1.0%)<sup>1</sup> | Low Response,<br>N = 25 928 (33.3%)<sup>1</sup> | Overall,<br>N = 77 957 (100%)<sup>1</sup> |
| --- | --- | --- | --- | --- | --- | --- |
| Class-membership Probability | 0.6 (0.5, 0.7) | 0.9 (0.6, 1.0) | 0.7 (0.5, 0.9) | 0.8 (0.6, 1.0) | 0.7 (0.6, 0.9) | 0.6 (0.5, 0.8) |
| Age at admission (years) | 69.2 (51.6, 81.1) | 68.4 (46.7, 81.7) | 70.3 (56.4, 81.0) | 70.1 (56.0, 81.6) | 63.6 (43.6, 79.3) | 67.8 (49.6, 80.6) |
| Sex (Male) | 18 □ 865 (52.3%) | 2 □ 240 (49.5%) | 6 □ 178 (57.9%) | 415 (55.9%) | 11 □ 363 (43.8%) | 39 □ 061 (50.1%) |
| Charlson score | 1 (0, 2) | 1 (0, 2) | 1 (1, 3) | 2 (1, 3) | 1 (0, 2) | 1 (0, 2) |
| Elixhauser score | 2 (1, 4) | 2 (1, 4) | 3 (2, 4) | 3 (2, 4) | 2 (1, 4) | 2 (1, 4) |
| Community-onset | 28 □ 184 (78.1%) | 4 □ 010 (88.5%) | 7 □ 162 (67.1%) | 521 (70.1%) | 22 □ 189 (85.6%) | 62 □ 066 (79.6%) |
| Immunosuppression | 4 □ 909 (13.6%) | 533 (11.8%) | 2 □ 222 (20.8%) | 164 (22.1%) | 3 □ 797 (14.6%) | 11 □ 625 (14.9%) |
| Diabetes mellitus | 7 □ 225 (20.0%) | 921 (20.3%) | 2 □ 335 (21.9%) | 158 (21.3%) | 4 □ 862 (18.8%) | 15 □ 501 (19.9%) |
| Palliative care | 1 □ 995 (5.5%) | 200 (4.4%) | 1 □ 332 (12.5%) | 79 (10.6%) | 892 (3.4%) | 4 □ 498 (5.8%) |
| >1 blood cultures in episode | 9 □ 386 (26.0%) | 1 □ 792 (39.6%) | 6 □ 096 (57.2%) | 495 (66.6%) | 4 □ 688 (18.1%) | 22 □ 457 (28.8%) |
| >1 positive blood cultures in episode | 706 (2.0%) | 136 (3.0%) | 663 (6.2%) | 27 (3.6%) | 191 (0.7%) | 1 □ 723 (2.2%) |
| Baseline antimicrobial susceptibility |  |  |  |  |  |  |
| Culture-negative | 30 □ 916 (85.7%) | 3 □ 550 (78.4%) | 8 □ 352 (78.3%) | 642 (86.4%) | 24 □ 004 (92.6%) | 67 □ 464 (86.5%) |
| Potential contaminant(s) | 1 □ 671 (4.6%) | 245 (5.4%) | 673 (6.3%) | 65 (8.7%) | 1 □ 274 (4.9%) | 3 □ 928 (5.0%) |
| Susceptible | 2 □ 943 (8.2%) | 637 (14.1%) | 1 □ 284 (12.0%) | 20 (2.7%) | 402 (1.6%) | 5 □ 286 (6.8%) |
| Resistant | 285 (0.8%) | 45 (1.0%) | 173 (1.6%) | 5 (0.7%) | 84 (0.3%) | 592 (0.8%) |
| No antimicrobial recorded | 84 (0.2%) | 21 (0.5%) | 43 (0.4%) | 3 (0.4%) | 63 (0.2%) | 214 (0.3%) |
| Unknown | 192 (0.5%) | 31 (0.7%) | 141 (1.3%) | 8 (1.1%) | 101 (0.4%) | 473 (0.6%) |

<sup>1</sup>Median (IQR); n (%)

In groups with peaks on day 1/2, CRP levels initially rose dramatically, then dropped and stabilised by day 8 (**Figure 2A**). The group peaking on day 2, however, had lower starting levels, potentially due to enrichment with community-onset infections (88.5% vs. 78.1%, SMD=0.28, **Table S3**). More of those peaking on day 2 also had >1 blood culture taken in their episode (39.6% vs. 26.0%, SMD=0.29, **Table S3**), and more had pathogens cultured (16.2%[734/4,529] vs. 9.7%[3,504/36,091], **Figure S9A**). The slow recovery group had the highest peak yet recovered the slowest. Compared with those peaking on day 1, this group were older (median 70.3 vs. 69.2 years, SMD=0.13), had more comorbidities, immunosuppression (20.8% vs. 13.6%, SMD=0.19), nosocomial infections (32.9% vs. 21.9%, SMD=0.25), >1 positive blood cultures in the episode (6.2% vs. 2.0%, SMD=0.22) and more resistant baseline antimicrobials (2.0% vs. 1.2%, SMD=0.2, **Table S3**). The very small group who peaked 6 days after the episode started had a similar profile to the slow recovery group, with even more comorbidities and episodes with >1 blood culture (66.6% vs. 26.0% in those peaking on day 1, SMD=0.89, **Table S3**). Mean CRP in the low response group remained <20mg/L throughout; this group had fewer blood cultures (18.1% with >1 blood culture vs. 26.0% in those peaking on day 1, SMD=0.19), more negative cultures (92.6% vs. 85.7%) and more community-onset infections (85.6% vs. 78.1%, SMD=0.20); patients were generally younger (median 63.6 vs. 69.2 years, SMD=0.19, **Table S3**).

Estimated response trajectories for heart rate, respiratory rate, temperature, and WBC count by the latent CRP trajectory class showed the same response patterns in terms of early/delayed/low response (**Figure S10**).

### Expected CRP response

To estimate the “normal” response to suspected BSI treated with effective antibiotics (either empirically or through prompt switching), and expected variation in this, regardless of whether a pathogen was identified, we included 40,620 episodes in the groups with CRP peaking on day 1/2, i.e., those who exhibited a “typical” response. Estimated centile charts based on 100,000 bootstrap samples (assuming that the observed episodes’ characteristics would generalise to the population presenting to the hospital with suspected BSI) show 5th, 10th, 25th, 50th, 75th, 90th, and 95th percentiles of a normal CRP response to suspected BSI, whether subsequently culture positive or not (**Figure 3A**). Estimates were similar when randomly sampling one measurement per patient, suggesting that potential for bias arising from multiple measurements was limited (**Figure S11**). Overall, the median level peaked at ∼165mg/L on day 1 (24h after blood culture collection), after which it decreased gradually to ∼25mg/L by day 8. This chart illustrates clearly the challenges on relying on absolute CRP value to determine response, or even change in CRP (**Figure 3B**), given individual-level heterogeneity. For example, a value of 150mg/L would be expected (50th percentile) 12h after blood culture collection for an average responder, but would still represent a standard (i.e. good) response at 2.7 days for a patient whose initial CRP was higher (75th percentile) and even later, at 3.7 and 4.2 days, for even higher initial CRP (90th and 95th percentile respectively). We also estimated the expected CRP response centiles (10th, 50th, 90th) for different sources of infection separately based on relevant episode subgroups, which showed little difference (**Figure S12**).

**Figure 3.**
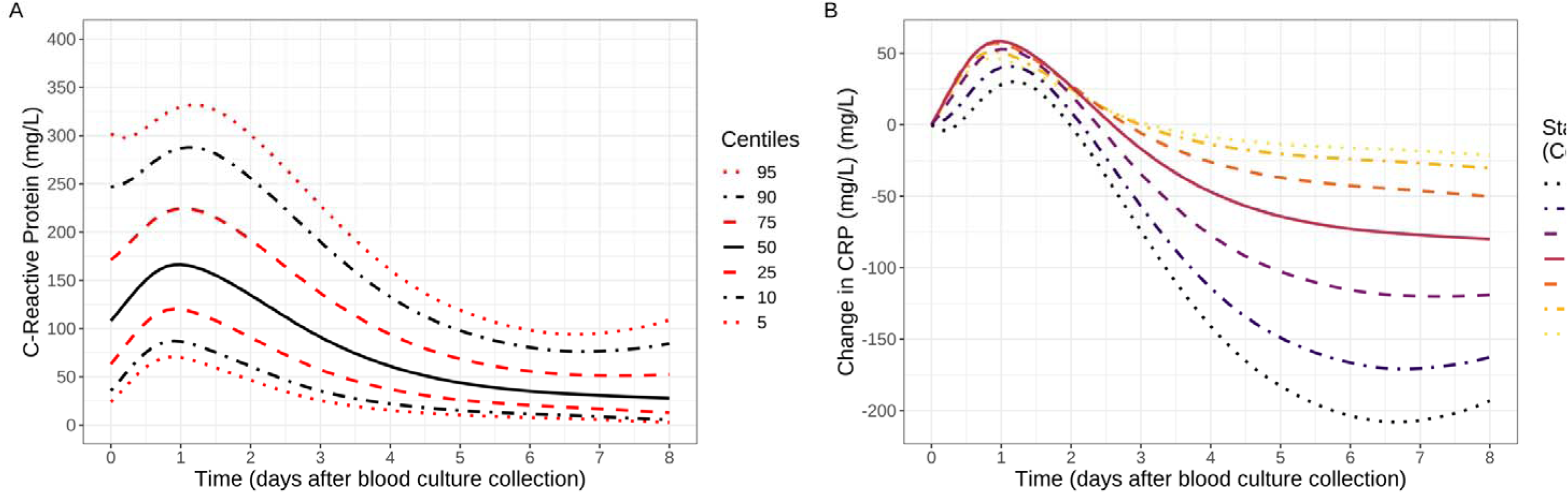
Centile reference chart of expected CRP response in patients with culture-positive/negative suspected BSI responding standardly to antibiotics (A) and change in CRP from initial value in centile (B). Change in CRP was calculated by subtracting the CRP value at the datetime of blood culture collection. Note: estimated from the two latent classes peaking on day 1 and 2 in Figure 2, regardless of pathogen isolated.

## Discussion

Using large-scale EHR data, we showed clinical response trajectories in laboratory tests and vital signs are associated with both specific blood culture results and sources of infection in patients with suspected BSI. We found considerable variation across different pathogen groups in response trajectories with much of the variation driven by differences around presentation. Five distinct patterns of CRP response trajectories were identified using latent class models, providing evidence for heterogeneity in infection responses; interestingly nearly 90% of culture-positive episodes with a pathogen, but also around two-thirds of culture-negative episodes, were associated with acute response. Centile reference charts were created based on the typical CRP responders to standardise assessment of infection progression and treatment response in patients with suspected BSI; these could be used to guide management independent of microbiological test results.

Response trajectories were strongly associated with sources of infection and culture results/pathogen groups, at least partly explaining heterogeneity in host responses to BSI. Higher and more prolonged inflammatory responses with abdominal and multiple sources of infection are consistent earlier studies,^21,22^ and associations with higher mortality.^21^ Although previous studies suggest Gram-negative infections generally cause stronger inflammatory responses (e.g., in PCT and CRP),^22,23^ we found a more pronounced CRP, WBC, heart rate, respiratory rate, and temperature responses to some Gram-positive bacteria (particularly *S. pneumoniae* and beta-haemolytic Streptococci) after adjusting for sources of infection. This may reflect analysing different Gram-positive infections separately, and our approach considering multiple measurements compared to most literature relying on single time-points. Infections susceptible to baseline antimicrobials were associated with higher CRP responses than resistant infections, possibly due to increased initial inflammatory responses from antimicrobial killing or a reduced fitness cost from antimicrobial resistance. However, this difference was not apparent in other physiological measurements.

We considered those in the groups with peak CRP levels on day 1/2 (52.1% of episodes) to have a “normal” response (assumed to also represent appropriate antibiotic treatment). The day 2 peak group, may represent a slightly delayed response or detection of suspected BSI earlier in the illness. The slow recovery group was characterised by stronger initial and more persistently elevated CRP and, like the small group with a delayed peak on day 6, included more older patients with more comorbidities. The group with limited CRP response included younger patients with more negative blood cultures, with mean estimates likely reduced by the absence of bacterial infection or a systemic response in a substantial subset.

Whilst CRP response trajectories have not been described in this detail to our knowledge, host response characteristics and clinical outcomes have previously been used to sub-phenotype patients with suspected BSI or sepsis. However, many studies used only baseline laboratory test results and physiological indicators or static measurements post-sepsis onset.^8,24^ Several studies studied longitudinal measures of vital signs, WBC or SOFA scores.^7,12–18^ Broadly mirroring our observations, three studies identified four temperature trajectory groups using measurements within the first 72h: “hyperthermic, slow resolvers”, “hyperthermic, fast resolvers”, “normothermic”, and “hypothermic”.^11–13^ For example, the group with CRP peaking on day 1/2 had temperature responses corresponding to the “hyperthermic, fast resolvers”; the late CRP response group likely corresponded to the “hypothermic” group, both comprising older patients with more comorbidities; however, although the slow recovery CRP group had a similar temperature trajectory to the “hyperthermic, slow resolvers”, our group consisted mainly of relatively old rather than young patients as previously. WBC response trajectories estimated by the latent CRP groups were also broadly consistent with previous study that identified seven white blood cell trajectories from 917 ICU patients.^15^

Despite the heterogeneity seen in CRP responses by pathogens and clinical syndromes, for a given starting value of CRP, responses were relatively consistent. This means that CRP responses could be summarised using a single centile reference chart. Although the same absolute value or even change in CRP means something different depending on where a patient’s CRP started, this can be accounted for. This heterogeneity in CRP response trajectories illustrates the limitations of a “one-size-fits-all” approach to using absolute CRP values to determine escalation, de-escalation or duration of antibiotic therapy in patients with suspected BSI, whereas the centile chart presented provides a potentially useful alternative. Spotting unexpected deterioration is key potential application and biomarker-guided antibiotic stewardship is another.^5,25^ Though previous biomarker guided stewardship reduced antibiotic prescription and duration while demonstrating non-inferior or lower mortality, compliance remained suboptimal.^26–28^ The centile reference chart provides a more visually intuitive means of assessing biomarker response, potentially aiding clinical decisions by incorporating individual-level observations alongside evidence-based references. Its implementation could be supported by embedding it within EHR systems.

Study strengths include our large sample size (77,957 suspected BSI episodes) and longer duration of follow up (8 days) compared to previous studies, as well as using comprehensive clinical data over several years. There are several limitations to our approach. As we use data collected for clinical reasons, and CRP and other laboratory measurements are less likely to be (serially) repeated in those making a good recovery, measurements at later time points are likely enriched for elevated values. Hence true expected trajectories may fall more rapidly and more completely than we estimate. Mitigating this entirely would require a design that sampled irrespective of clinical progress and post-discharge, which is unlikely to be feasible at the scale of our study. Other limitations include the fact that no CRP measurement was available in 10,391 (11.8%) episodes with suspected BSI. Our relatively high culture-negative rate (87.3%) is partly due to our broad definition of suspected infection and historically high rate of taking blood cultures; nevertheless 51.1% culture-negative episodes still exhibited typical CRP responses, peaking on day 1/2. Only the association between baseline antimicrobial treatment activity and CRP response was examined; future planned work includes investigating associations between CRP levels/centiles and changes in antimicrobial therapy, both to assess if there is evidence that sub-optimal responses in CRP and other markers lead to changes in antimicrobials and also if switching from inactive to active therapy changes CRP trajectories. PCT can also help guide the duration of antibiotic therapy, but this biomarker was not measured routinely at our hospitals. Despite using bootstrapping and simulations, EHR data may contain inaccuracies or missing information, potentially impacting the estimation of clinical response trajectories. Furthermore, our analysis was limited to patient data available in one, albeit large, hospital group, which might influence the generalisability of our findings.

## Conclusions

Our analysis revealed strong associations between clinical response trajectories and both sources of infection and different pathogen groups in patients with suspected BSI, with distinct CRP response patterns, reflecting normal, slow, and delayed or limited responses. Considering the dynamic nature of BSI and sepsis and heterogeneity in individual CRP response trajectories, the centile reference charts developed in this study may provide a valuable tool for guiding individualised infection management. Future research should focus on exploring the dynamic association between response and antibiotic use and evaluating the practical application of centile reference charts in clinical settings.

## Supporting information

see supplement

## Data Availability

The data analysed are available from the Infections in Oxfordshire Research Database (https://oxfordbrc.nihr.ac.uk/research-themes/modernising-medical-microbiology-and-big-infection-diagnostics/infections-in-oxfordshire-research-database-iord/), subject to an application and research proposal meeting on the ethical and governance requirements of the Database.

## Acknowledgements and funding

This work was supported by the National Institute for Health Research Health Protection Research Unit (NIHR HPRU) in Healthcare Associated Infections and Antimicrobial Resistance at Oxford University in partnership with the UK Health Security Agency (NIHR200915), and the NIHR Biomedical Research Centre, Oxford. DWE is a Big Data Institute Robertson Fellow. ASW is an NIHR Senior Investigator. The views expressed are those of the authors and not necessarily those of the NHS, the NIHR, the Department of Health or the UK Health Security Agency. The funders had no role in study design, data collection and analysis, decision to publish, or preparation of the manuscript.

