## Supplementary material for "Defining normal inflammatory marker and vital sign responses to suspected bloodstream infection in adults with positive and negative blood cultures": see supplement

### Supplementary Methods

Linear mixed models were used to estimate CRP, WBC and vital signs’ trajectories throughout suspected BSI episodes, from -1 day (CRP, WBC) or -6 hours (vital signs) before to +8 days after the start of each episode, using box-cox transformed values as the outcome. Nonlinear trends were incorporated via natural cubic splines with four knots at the 20th, 40th, 60th and 80th percentiles of observed time values and included as fixed and random effects, as was the intercept (i.e. vital sign/laboratory test value at the start of each episode). Fixed effects were included for source of infection, community-onset (blood sample collection ≤48 hours from admission), blood culture result (positive, potential contaminant, negative) and pathogen group (grouped as in **Table S1**), age, sex, Charlson comorbidity and Elixhauser acuity scores, and immunosuppression. Charlson and Elixhauser scores were calculated using ICD-10 diagnostic codes based on a 1-year lookback period^1^. Immunosuppression was determined by the presence of ICD-10 diagnostic codes for AIDS/HIV, metastatic cancer, haematological malignancies, primary immunodeficiencies and end-stage liver disease within the same 1-year lookback period. These main effects reflected associations with the value at the start of each episode; we tested interactions between each covariate and the natural cubic splines for time and included those with p<0.05, i.e. those with an impact on the trajectory as well as the baseline values (blood culture result and pathogen group, infection source, community-onset and immunosuppression).

Separate adjusted models were fitted to examine effects of source of infection and baseline antimicrobial susceptibility. Baseline antimicrobials were those administered within 12h before to 24h after blood culture collection. The baseline antimicrobial susceptibility profile for each suspected BSIs episode was evaluated based on the results of antimicrobial susceptibility testing as well as information on intrinsic resistance and antimicrobial activity in the Sanford Guide if susceptibility test results were not available.^2^ Where results were not available and there was uncertainty about the expected susceptibility then results were recorded as unknown. The susceptibility profile of rarer pathogens were defined as unknown where these were not tested and intrinsic susceptibility data were not available. Episodes in the intensive care unit, where antimicrobial administration was recorded using a different EHR system from which data were not available (39 cases), were also defined as having unknown susceptibility. For polymicrobial infections, resistance of any one pathogen to the baseline antibiotics defined the episode as antibiotic-resistant. Candida infections treated with only baseline antibiotics (98.6%[73/74]) were also defined as resistant to baseline therapy.

Unadjusted latent class mixed models (LCMM) were used to identify underlying population-level heterogeneity in the response trajectories of routinely collected CRP measurements. We fitted LCMMs with between 1 and 6 classes, and the optimal number of classes was chosen based on the Bayesian Information Criterion (BIC) and percentage of class membership (≥0.5%) (**Table S4**). Patient characteristics and other covariates were not included in the models because we did not want to adjust for these potential causes of underlying heterogeneity. From the selected LCMM, each episode was assigned to the class with the highest posterior probability. We then compared characteristics between the latent class groups univariably. The Standardised Mean Difference (SMD) was used in the comparison to maintain consistency and to better account for differences in measurement scales and for an improved reflection of effect sizes across our study variables. We did not employ the three-step approach to test class membership predictors as the package we used (lcmm 2.0.0) did not support this methodology yet.^3,4^

Centile reference charts for expected CRP response in standard responders (i.e. those with peak response on day 1–2) were constructed using the lambda-mu-sigma (LMS) method^5^, adopted by the World Health Organization (WHO) to generate the childhood growth standards^6^, and implemented using Generalised Additive Models for Location Scale and Shape library (GAMLSS, version 5.4.3)^7^. Three candidate distributions were considered: Box-Cox Cole Green (truncated standard normal distribution), Box-Cox power exponential (truncated exponential power distribution), and Box-Cox t (truncated t distribution), selected using the Generalised Akaike Information Criterion^8^, with degrees of freedom for the penalised spline model smoothing parameters determined using the local Schwarz Bayesian Criterion^8^. With these parameters, z-scores and percentiles of CRP response can be generated for times after blood culture collection.

The LMS method assumes the measurements used for constructing centile charts are independent, whereas serial measurements of vital signs and laboratory tests are correlated within individuals^9^. This could potentially cause bias, especially if patients with abnormal responses get prolonged and more frequent measurements. The common solution of selecting one random observation for each patient can result in significant information loss. We addressed the problem of potentially informative numbers of measurements by constructing 100,000 bootstrap samples of the 40,620 episodes from standard responders and using a linear mixed model with only fixed and random effects for time (natural cubic spline) as above to simulate the CRP values of these patients at nine random time points up to 8 days after the start of each episode (incorporating fixed effects, correlated random effects per patient and additional measurement error in the prediction) then using these values (100,000 episodes, 900,000 observations) as outcomes for the LMS model.

Analyses were performed using statistical software R, version 4.1.0 (R Project for Statistical Computing).

### Supplementary Tables

| **Blood Culture Results** | **CRP Measured**, N = 77 957 (88.2%)^1^ | **CRP Not Measured**, N = 10 391 (11.8%)^1^ | **Difference**^2^ |
| --- | --- | --- | --- |
| Gram-positive Pathogens | | | |
| *Staphylococcus aureus* | 697 (0·9%) | 19 (0·2%) | 0·10 |
| Beta-Hemolytic Streptococci | 389 (0·5%) | 32 (0·3%) | 0·03 |
| *Enterococcus* sp. | 329 (0·4%) | 14 (0·1%) | 0·05 |
| *Streptococcus pneumoniae* | 290 (0·4%) | 16 (0·2%) | 0·04 |
| Other Pathogenic *Streptococcus* | 124 (0·2%) | 4 (0·0%) | 0·04 |
| Gram-negative Pathogens | | | |
| *Escherichia coli* | 2 284 (2·9%) | 126 (1·2%) | 0·12 |
| *Klebsiella* sp. | 503 (0·6%) | 21 (0·2%) | 0·07 |
| Other *Enterobacterales* | 349 (0·4%) | 18 (0·2%) | 0·05 |
| *Pseudomonas aeruginosa* | 289 (0·4%) | 14 (0·1%) | 0·05 |
| *Enterobacter* sp. | 127 (0·2%) | 5 (0·0%) | 0·04 |
| Other Pathogens | | | |
| Other | 510 (0·7%) | 45 (0·4%) | 0·03 |
| Polymicrobial | 309 (0·4%) | 6 (0·1%) | 0·07 |
| Anaerobes | 284 (0·4%) | 23 (0·2%) | 0·03 |
| *Candida* sp. | 81 (0·1%) | 2 (0·0%) | 0·03 |
| Potential Contaminant(s) and Culture-negative | | | |
| Culture-negative | 67 464 (86·5%) | 9 667 (93·0%) | 0·22 |
| CoNS (contaminant) | 3 184 (4·1%) | 305 (2·9%) | 0·06 |
| Other Suspected Contaminants | 441 (0·6%) | 59 (0·6%) | 0·00 |
| Viridans and Other *Streptococcus* | 303 (0·4%) | 15 (0·1%) | 0·05 |
| ^1^n (%) | | | |
| ^2^Standardised Mean Difference | | | |

**Table S1.** Comparison of blood culture results for suspected BSIs episodes with ≥1 measurement of CRP within 1 day before to 8 days after the start of each episode versus those without any CRP measurements. Other Pathogenic *Streptococcus* includes *Streptococcus anginosus*, *Streptococcus gallolyticus*, *Streptococcus constellatus*, *Streptococcus intermedius*, *Streptococcus lutetiensis*, *Streptococcus bovis*. CoNS (contaminant) refers to Coagulase negative staphylococci. Percentages in the header are of all episodes, and in the main body are column percentages within each group. Effect size estimated using standardised mean difference (SMD), considering 0.2, 0.5, and 0.8 as small, medium, and large, respectively.

|  | Baseline Antimicrobial Susceptibility | | | | |
| --- | --- | --- | --- | --- | --- |
|  | Susceptible | Resistant | No Antimicrobial Recorded | Unknown | Total |
| Gram-positive Pathogens | | | | | |
| *Staphylococcus aureus* | 606 (87%) | 21 (3·0%) | 34 (4·9%) | 36 (5·2%) | 697 (100%) |
| Beta-Hemolytic Streptococci | 373 (96%) | 0 (0%) | 13 (3·3%) | 3 (0·8%) | 389 (100%) |
| *Enterococcus* sp. | 228 (69%) | 49 (15%) | 12 (3·6%) | 40 (12.2%) | 329 (100%) |
| *Streptococcus pneumoniae* | 278 (96%) | 0 (0%) | 9 (3·1%) | 3 (1·0%) | 290 (100%) |
| Other Pathogenic *Streptococcus* | 101 (81%) | 2 (1·6%) | 16 (13%) | 5 (4·0%) | 124 (100%) |
| Gram-negative Pathogens | | | | | |
| *Escherichia coli* | 2 127 (93%) | 88 (3·9%) | 35 (1·5%) | 34 (1·5%) | 2 284 (100%) |
| *Klebsiella* sp. | 445 (88%) | 20 (4·0%) | 9 (1·8%) | 29 (5·8%) | 503 (100%) |
| Other *Enterobacterales* | 281 (81%) | 40 (11%) | 7 (2·0%) | 21 (6·0%) | 349 (100%) |
| *Pseudomonas aeruginosa* | 185 (64%) | 74 (26%) | 7 (2·4%) | 23 (8·0%) | 289 (100%) |
| *Enterobacter* sp. | 74 (58%) | 44 (35%) | 4 (3·1%) | 5 (3·9%) | 127 (100%) |
| Other Pathogens | | | | | |
| Other | 183 (36%) | 64 (13%) | 48 (9·4%) | 215 (42.2%) | 510 (100%) |
| Polymicrobial | 175 (57%) | 88 (28%) | 6 (1·9%) | 40 (12.9%) | 309 (100%) |
| Anaerobes | 226 (80%) | 28 (9·9%) | 12 (4·2%) | 18 (6·3%) | 284 (100%) |
| *Candida* sp. | 4 (4·9%) | 74 (91%) | 2 (2·5%) | 1 (1·2%) | 81 (100%) |
| Total | 5 286 (81%) | 592 (9·0%) | 214 (3·3%) | 473 (7·2%) | 6 565 (100%) |

**Table S2.** Distribution of baseline antimicrobial susceptibility across pathogen groups for suspected BSI episodes with ≥1 measurement of CRP within 1 day before to 8 days after the start of each episode. Culture-negative episodes and episodes with potential contaminants were excluded. See supplemental methods for definition of baseline antimicrobial susceptibility. Note: Baseline antimicrobials include both antibiotics and antifungals.

|  |  | | **Comparisons with Peak on Day 1** | | | | | | | |
| --- | --- | --- | --- | --- | --- | --- | --- | --- | --- | --- |
| **Characteristic** | | **Peak on Day 1** | **Peak on Day 2** | **Difference**^2^ | **Slow Recovery** | **Difference**^2^ | **Peak on Day 6** | **Difference**^2^ | **Low Response** | **Difference**^2^ |
| Age at admission (years) | | 69·2 (51·6, 81·1) | 68·4 (46·7, 81·7) | 0·06 | 70·3 (56·4, 81·0) | 0·13 | 70·1 (56·0, 81·6) | 0·11 | 63·6 (43·6, 79·3) | 0·19 |
| Sex (Male) | | 18 865 (52·3%) | 2 240 (49·5%) | 0·06 | 6 178 (57·9%) | 0·11 | 415 (55·9%) | 0·07 | 11 363 (43·8%) | 0·17 |
| Charlson score | | 1 (0, 2) | 1 (0, 2) | 0·01 | 1 (1, 3) | 0·16 | 2 (1, 3) | 0·20 | 1 (0, 2) | 0·04 |
| Elixhauser score | | 2 (1, 4) | 2 (1, 4) | 0·01 | 3 (2, 4) | 0·20 | 3 (2, 4) | 0·22 | 2 (1, 4) | 0·04 |
| Community-onset | | 28 184 (78·1%) | 4 010 (88·5%) | 0·28 | 7 162 (67·1%) | 0·25 | 521 (70·1%) | 0·18 | 22 189 (85·6%) | 0·20 |
| Immunosuppression | | 4 909 (13·6%) | 533 (11·8%) | 0·06 | 2 222 (20·8%) | 0·19 | 164 (22·1%) | 0·22 | 3 797 (14·6%) | 0·03 |
| Diabetes mellitus | | 7 225 (20·0%) | 921 (20·3%) | 0·01 | 2 335 (21·9%) | 0·05 | 158 (21·3%) | 0·03 | 4 862 (18·8%) | 0·03 |
| Palliative care | | 1 995 (5·5%) | 200 (4·4%) | 0·05 | 1 332 (12·5%) | 0·24 | 79 (10·6%) | 0·19 | 892 (3·4%) | 0·10 |
| >1 blood cultures in episode | | 9 386 (26·0%) | 1 792 (39·6%) | 0·29 | 6 096 (57·2%) | 0·67 | 495 (66·6%) | 0·89 | 4 688 (18·1%) | 0·19 |
| >1 positive blood cultures in episode | | 706 (2·0%) | 136 (3·0%) | 0·07 | 663 (6·2%) | 0·22 | 27 (3·6%) | 0·10 | 191 (0·7%) | 0·11 |
| Baseline antibiotic susceptibility | |  |  | 0·20 |  | 0·20 |  | 0·30 |  | 0·32 |
| Culture-negative | | 30 916 (85·7%) | 3 550 (78·4%) |  | 8 352 (78·3%) |  | 642 (86·4%) |  | 24 004 (92·6%) |  |
| Potential contaminant(s) | | 1 671 (4·6%) | 245 (5·4%) |  | 673 (6·3%) |  | 65 (8·7%) |  | 1 274 (4·9%) |  |
| Susceptible | | 2 943 (8·2%) | 637 (14·1%) |  | 1 284 (12·0%) |  | 20 (2·7%) |  | 402 (1·6%) |  |
| Resistant | | 285 (0·8%) | 45 (1·0%) |  | 173 (1·6%) |  | 5 (0·7%) |  | 84 (0·3%) |  |
| No antimicrobial recorded | | 84 (0·2%) | 21 (0·5%) |  | 43 (0·4%) |  | 3 (0·4%) |  | 63 (0·2%) |  |
| Unknown | | 192 (0·5%) | 31 (0·7%) |  | 141 (1·3%) |  | 8 (1·1%) |  | 101 (0·4%) |  |
| ^1^Median (IQR); n (%); ^2^Standardised Mean Difference | | | | | | | | | | |

**Table S3.** Comparison of episode characteristics between those estimated from latent class models as Peak on Day 2, Slow Recovery, Peak on Day 6, Low Response, and Peak on Day 1 as the reference group. Unadjusted effect size estimated using standardised mean difference (SMD), considering 0.2, 0.5, and 0.8 as small, medium, and large, respectively.

| **N classes** | **Log likelihood** | **AIC** | **BIC** | **SABIC** | **Entropy** | **Class 1 (%)** | **Class 2 (%)** | **Class 3 (%)** | **Class 4 (%)** | **Class 5 (%)** | **Class 6 (%)** |
| --- | --- | --- | --- | --- | --- | --- | --- | --- | --- | --- | --- |
| 1 | -522381.4 | 1044804.9 | 1044999.4 | 1044932.7 | 1.000 | 100.0 |  |  |  |  |  |
| 2 | -505915.1 | 1011884.3 | 1012134.4 | 1012048.6 | 0.856 | 0.7 | 99.3 |  |  |  |  |
| 3 | -499823.5 | 999713.0 | 1000018.7 | 999913.8 | 0.598 | 5.5 | 61.2 | 33.2 |  |  |  |
| 4 | -499309.0 | 998695.9 | 999057.2 | 998933.3 | 0.645 | 0.5 | 32.6 | 61.3 | 5.6 |  |  |
| 5 | -499012.2 | 998114.3 | 998531.2 | 998388.2 | 0.505 | 1.0 | 13.7 | 46.3 | 33.3 | 5.8 |  |
| 6 | -498126.1 | 996354.2 | 996826.7 | 996664.6 | 0.542 | 0.4 | 7.1 | 37.7 | 25.9 | 25.9 | 3.0 |

**Table S4.** Fit indices for latent class mixed models with different numbers of classes. Abbreviations: AIC: Akaike Information Criterion; BIC: Bayesian Information Criterion; SABIC: Sample-size adjusted BIC.

### Supplementary Figures

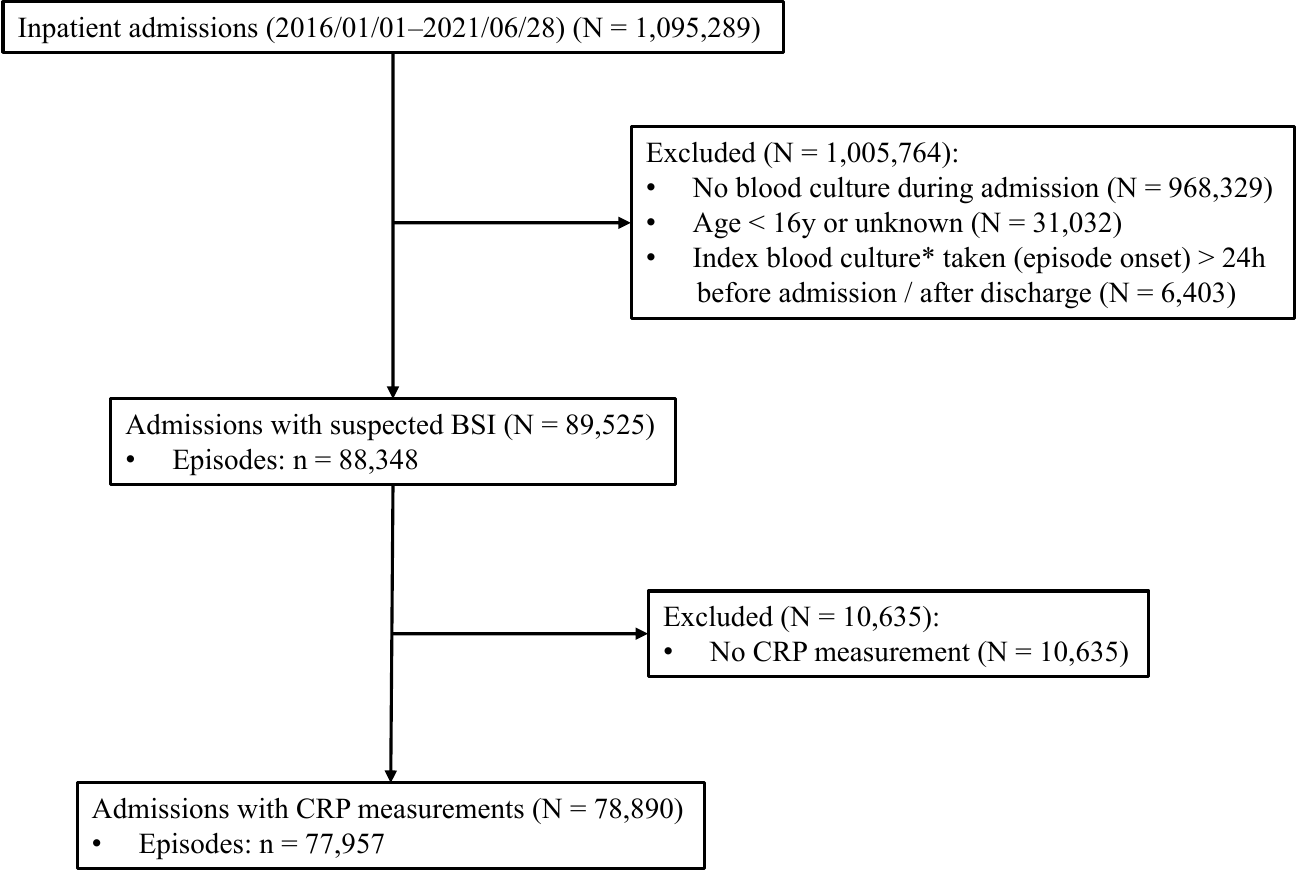

**Figure S1.** Flowchart for identifying suspected BSI infections and selecting the dataset for outcome investigations. *Index blood culture: the first blood sample from which a microbial pathogen was cultured, otherwise the first blood sample from which a contaminant was identified, or otherwise the first blood sample (if all cultures were negative). Note: For those excluded due to timing of index blood cultures, 70.0%[4,468/6,403] were culture-negative samples taken >24h before admission.

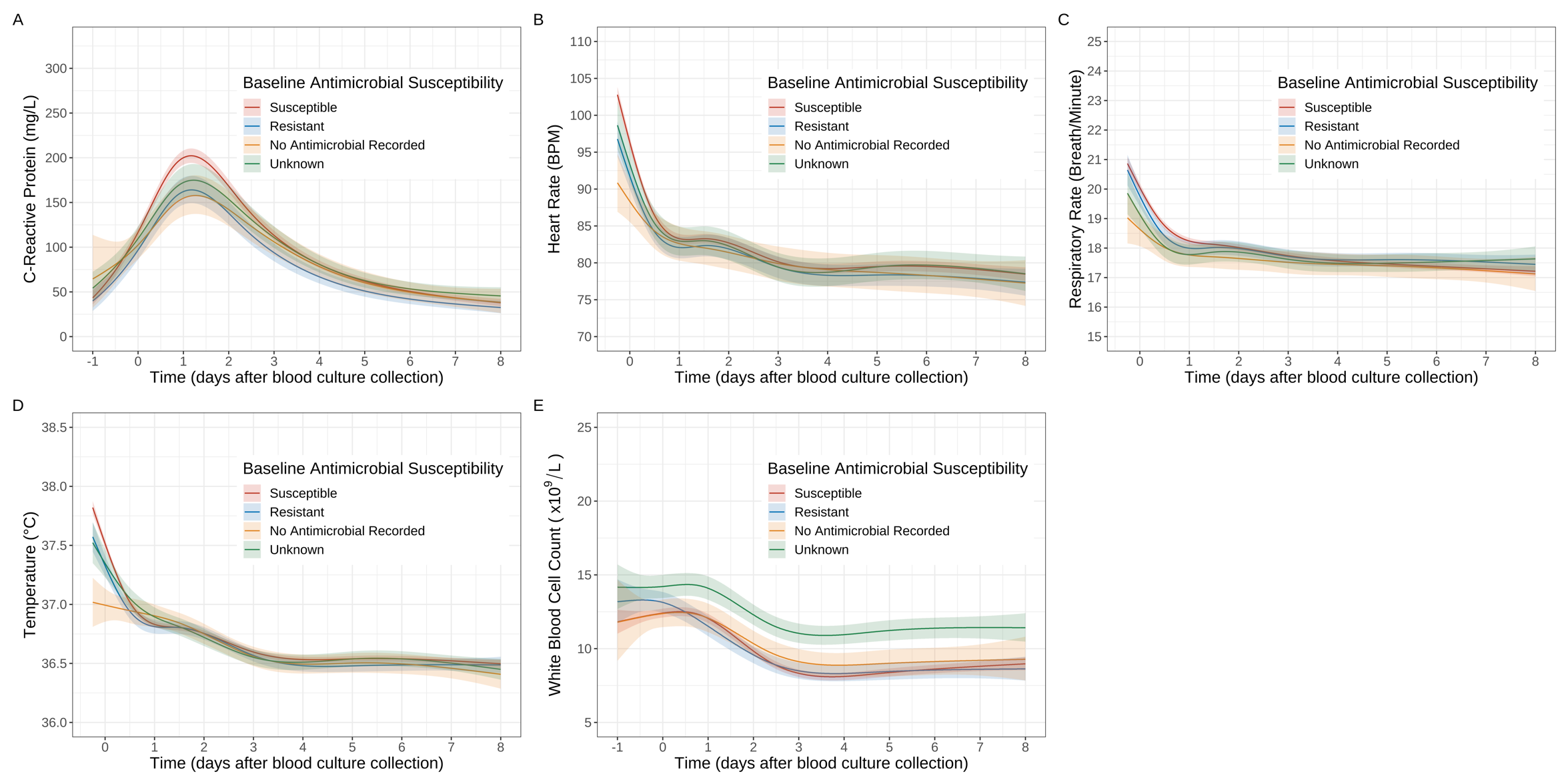

**Figure S2.** Response trajectories of CRP (A), heart rate (B), respiratory rate (C), body temperature (D) and WBC count (E) following different baseline antimicrobial susceptibilities. Predictions are plotted at the reference values of other adjusting variables: age = 64 years, male, Charlson score = 1, Elixhauser score = 3, community-onset, absence of immunosuppression, urinary source, and *E. coli* infection. Note: the no antimicrobial recorded group were enriched for people who were sufficiently well that they might not have received antimicrobials at baseline.

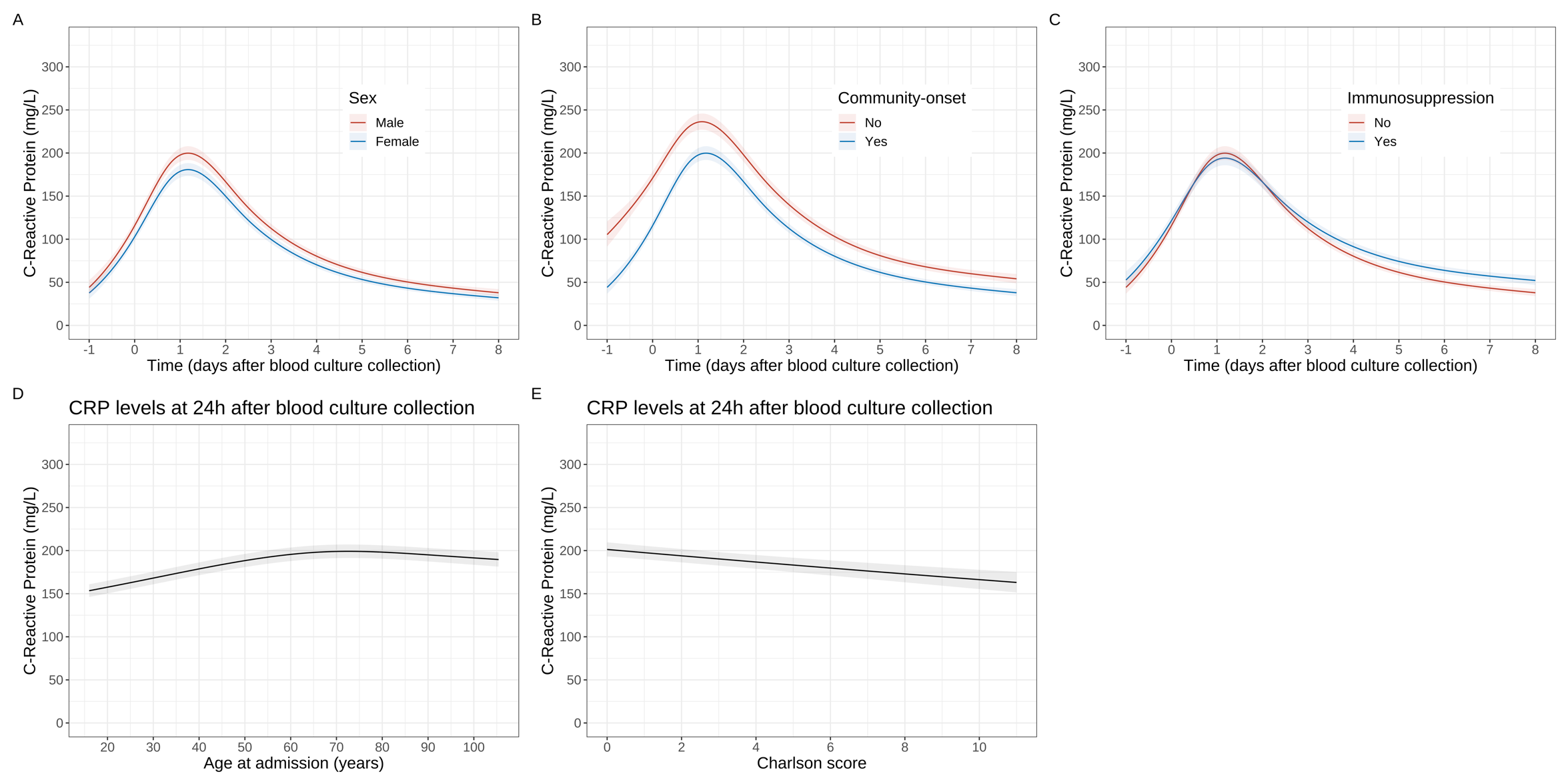

**Figure S3.** The adjusted associations between CRP response and sex (A), community-onset (B), immunosuppression (C), age (D) and Charlson score (E). Panels A–C present the relationships between CRP response trajectories from 1 days before to 8 days after the start of each episode. Panels D–E display CRP levels at 24 hours after the start of each episode to demonstrate the association between these continuous covariates and peak CRP levels (no evidence of interaction between these covariates and time; no evidence of association between Elixhauser and peak levels). Predictions are plotted at the reference values of other variables: age = 64 years, male, Charlson score = 1, Elixhauser score = 3, community-onset, absence of immunosuppression, urinary source, and *E. coli* infection. Nonlinear trends were incorporated via natural cubic splines with four knots at the 20th, 40th, 60th and 80th percentiles of observed time values (day 0, day 0.8, day 2.4, day 4.7).

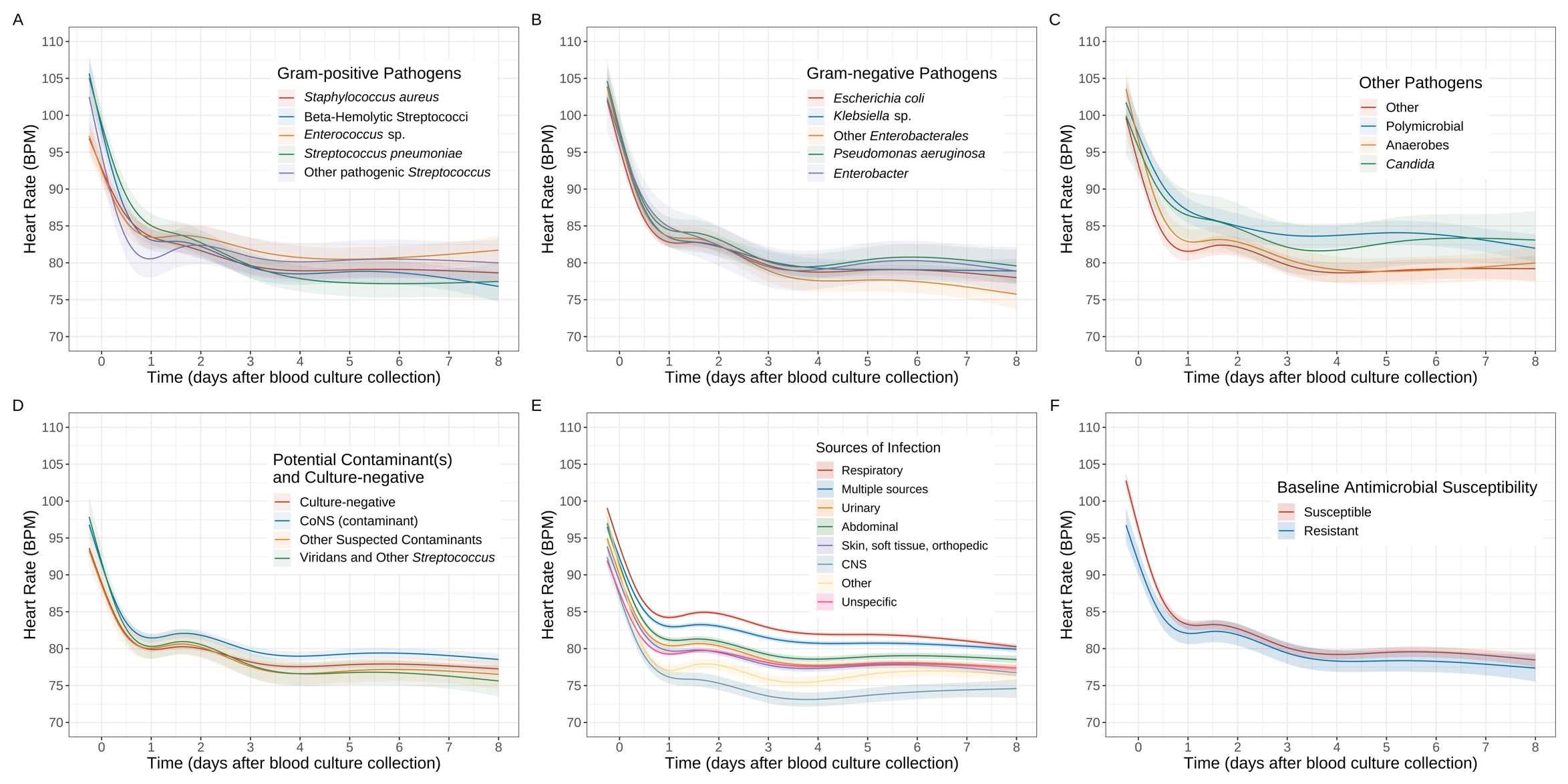

**Figure S4.** Heart rate response trajectories following different blood culture results (Gram-positive pathogens (A), Gram-negative pathogens (B), other pathogens (C), and potential contaminants and culture-negative results (D); adjusted for source of infection and other covariates), sources of infection (E) (not adjusted for blood culture results but adjusted for other covariates) and baseline antimicrobial susceptibilities (F) (adjusted for blood culture results, source of infection and other covariates). See **Figure S2B** for response trajectories of no baseline antimicrobial recorded and unknown baseline susceptibility. Predictions are plotted at the reference values of other adjusting variables: age = 64 years, male, Charlson score = 1, Elixhauser score = 3, community-onset, absence of immunosuppression, urinary source (excluding panel E), and *E. coli* infection (panel F only). Modelling time was limited to 6 hours prior to the start of each episode, as vital signs were not frequently measured before this point. Nonlinear trends were incorporated via natural cubic splines with four knots at the 20th, 40th, 60th and 80th percentiles of observed time values (day 0.4, day 1.5, day 3.0, day 5.1).

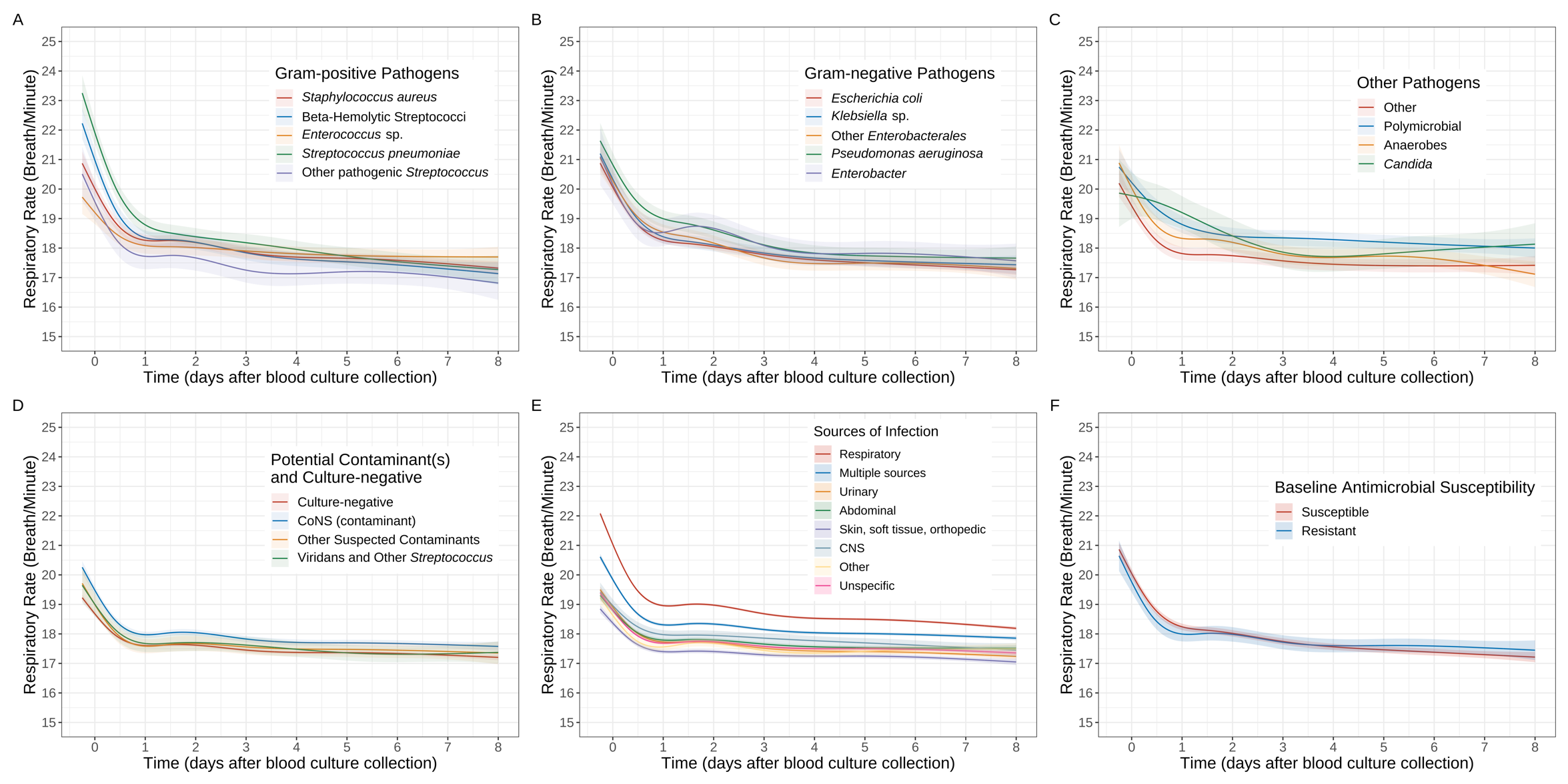

**Figure S5.** Respiratory rate response trajectories following different blood culture results (Gram-positive pathogens (A), Gram-negative pathogens (B), other pathogens (C), and potential contaminants and culture-negative results (D); adjusted for source of infection and other covariates), sources of infection (E) (not adjusted for blood culture results but adjusted for other covariates) and baseline antimicrobial susceptibilities (F) (adjusted for blood culture results, source of infection and other covariates). See **Figure S2C** for response trajectories of no baseline antimicrobial recorded and unknown baseline susceptibility. Predictions are plotted at the reference values of other adjusting variables: age = 64 years, male, Charlson score = 1, Elixhauser score = 3, community-onset, absence of immunosuppression, urinary source (excluding panel E), and *E. coli* infection (panel F only). Modelling time was limited to 6 hours prior to the start of each episode, as vital signs were not frequently measured before this point. Nonlinear trends were incorporated via natural cubic splines with four knots at the 20th, 40th, 60th and 80th percentiles of observed time values (day 0.4, day 1.5, day 3.0, day 5.1).

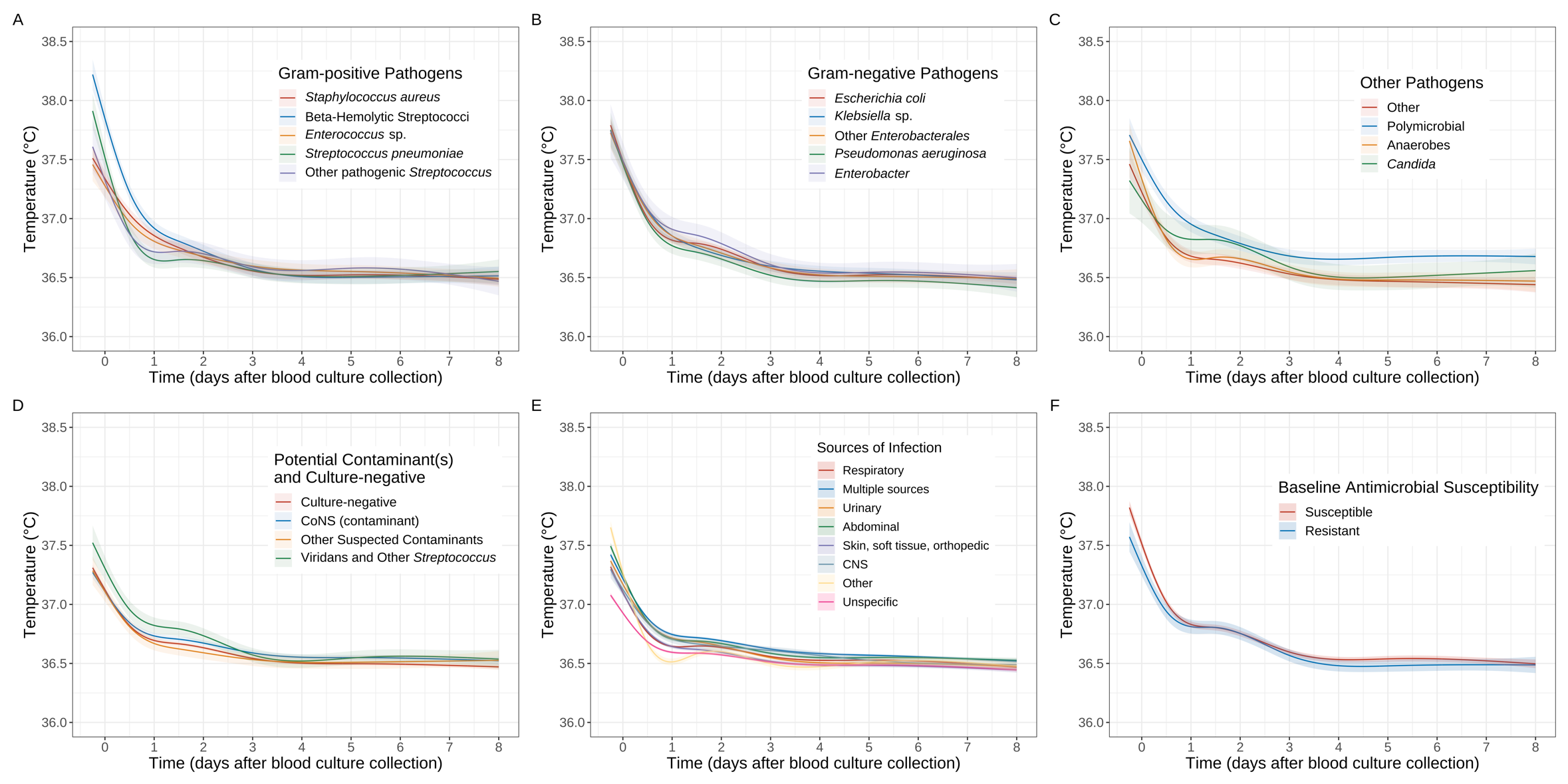

**Figure S6.** Body temperature response trajectories following different blood culture results (Gram-positive pathogens (A), Gram-negative pathogens (B), other pathogens (C), and potential contaminants and culture-negative results (D); adjusted for source of infection and other covariates), sources of infection (E) (not adjusted for blood culture results but adjusted for other covariates) and baseline antimicrobial susceptibilities (F) (adjusted for blood culture results, source of infection and other covariates). See **Figure S2D** for response trajectories of no baseline antimicrobial recorded and unknown baseline susceptibility. The acute temperature response with “other” source of infection in panel E (bright yellow) was potentially driven by brisk immune responses in younger patients with ENT and obstetric infections (and/or potential overfitting in this relatively small group, **Table 1**). Predictions are plotted at the reference values of other adjusting variables: age = 64 years, male, Charlson score = 1, Elixhauser score = 3, community-onset, absence of immunosuppression, urinary source (excluding panel E), and *E. coli* infection (panel F only). Modelling time was limited to 6 hours prior to the start of each episode, as vital signs were not frequently measured before this point. Nonlinear trends were incorporated via natural cubic splines with four knots at the 20th, 40th, 60th and 80th percentiles of observed time values (day 0.5, day 1.5, day 3.0, day 5.1).

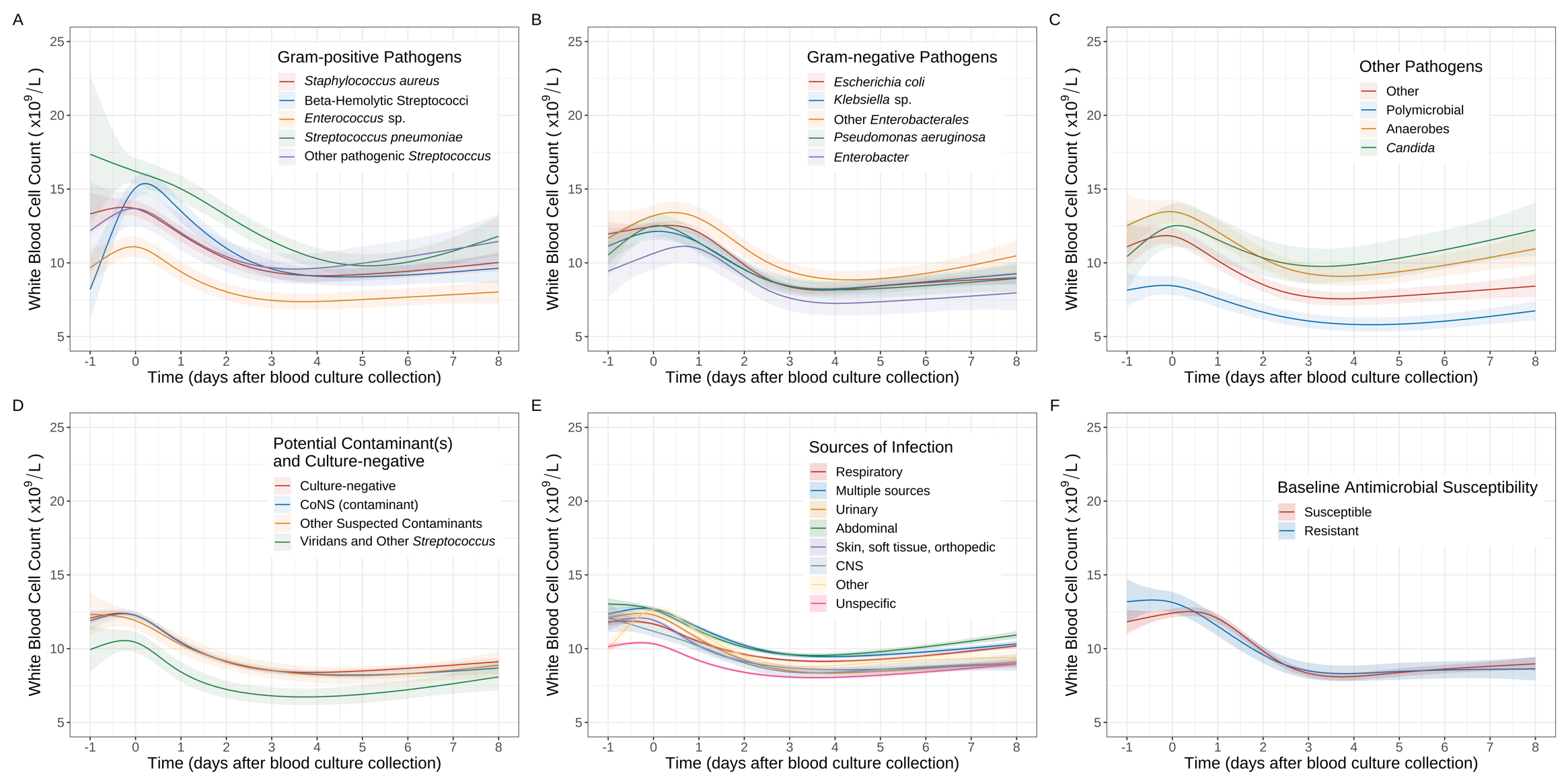

**Figure S7.** WBC count response trajectories following different blood culture results (Gram-positive pathogens (A), Gram-negative pathogens (B), other pathogens (C), and potential contaminants and culture-negative results (D); adjusted for source of infection and other covariates), sources of infection (E) (not adjusted for blood culture results but adjusted for other covariates) and baseline antimicrobial susceptibilities (F) (adjusted for blood culture results, source of infection and other covariates). See **Figure S2E** for response trajectories of no baseline antimicrobial recorded and unknown baseline susceptibility. The acute WBC count response with “other” source of infection in panel E (bright yellow) was potentially driven by brisk immune responses in younger patients with ENT and obstetric infections (and/or potential overfitting in this relatively small group, **Table 1**). Predictions are plotted at the reference values of other adjusting variables: age = 64 years, male, Charlson score = 1, Elixhauser score = 3, community-onset, absence of immunosuppression, urinary source (excluding panel E), and *E. coli* infection (panel F only). Nonlinear trends were incorporated via natural cubic splines with four knots at the 20th, 40th, 60th and 80th percentiles of observed time values (day 0, day 0.8, day 2.4, day 4.7).

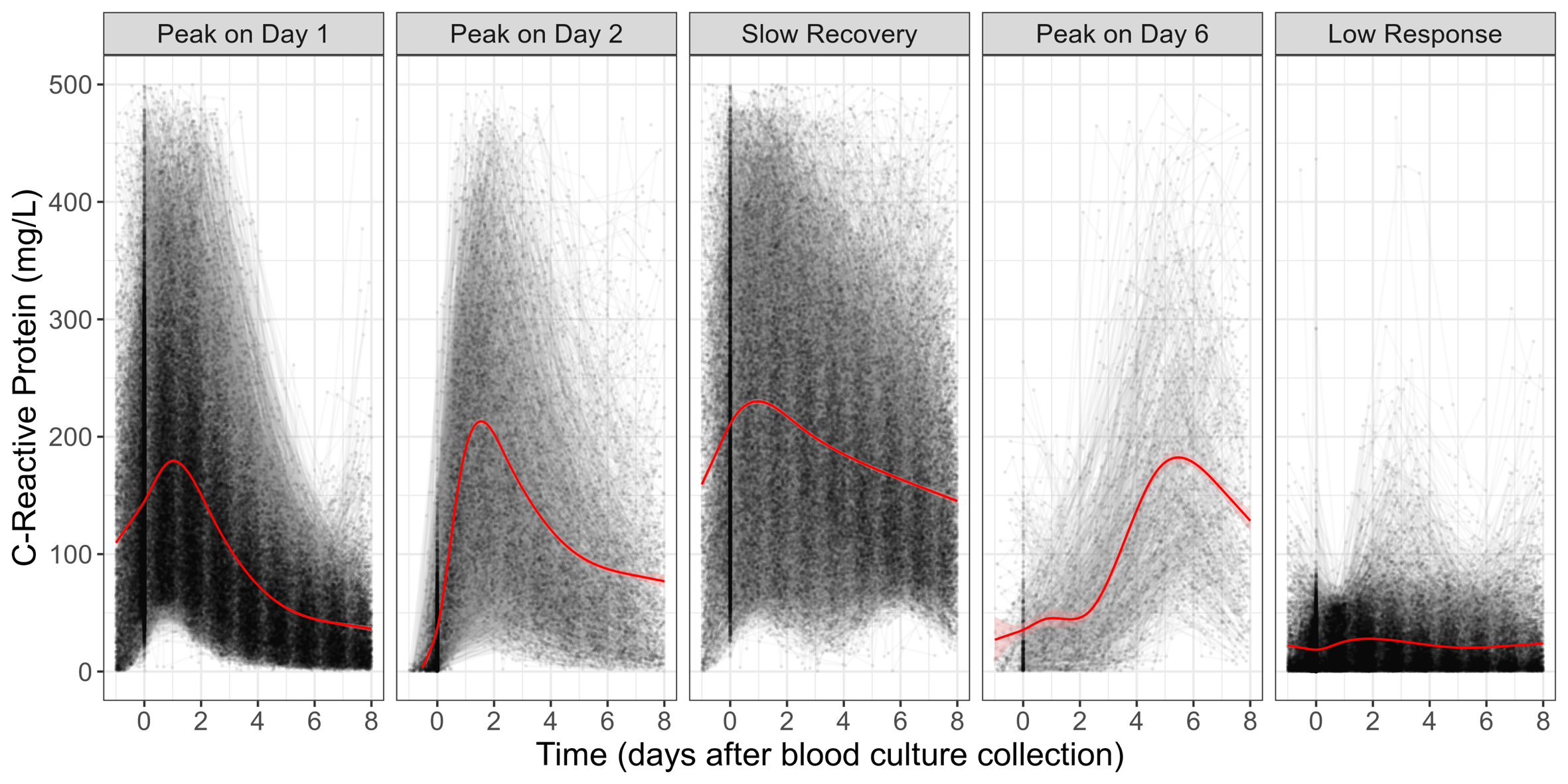

**Figure S8.** Spaghetti plot illustrating raw data underpinning latent classes of CRP response trajectories. Black lines represent individual CRP response trajectories for suspected BSIs episodes, while red lines show the mean response trajectory for each latent trajectory class.

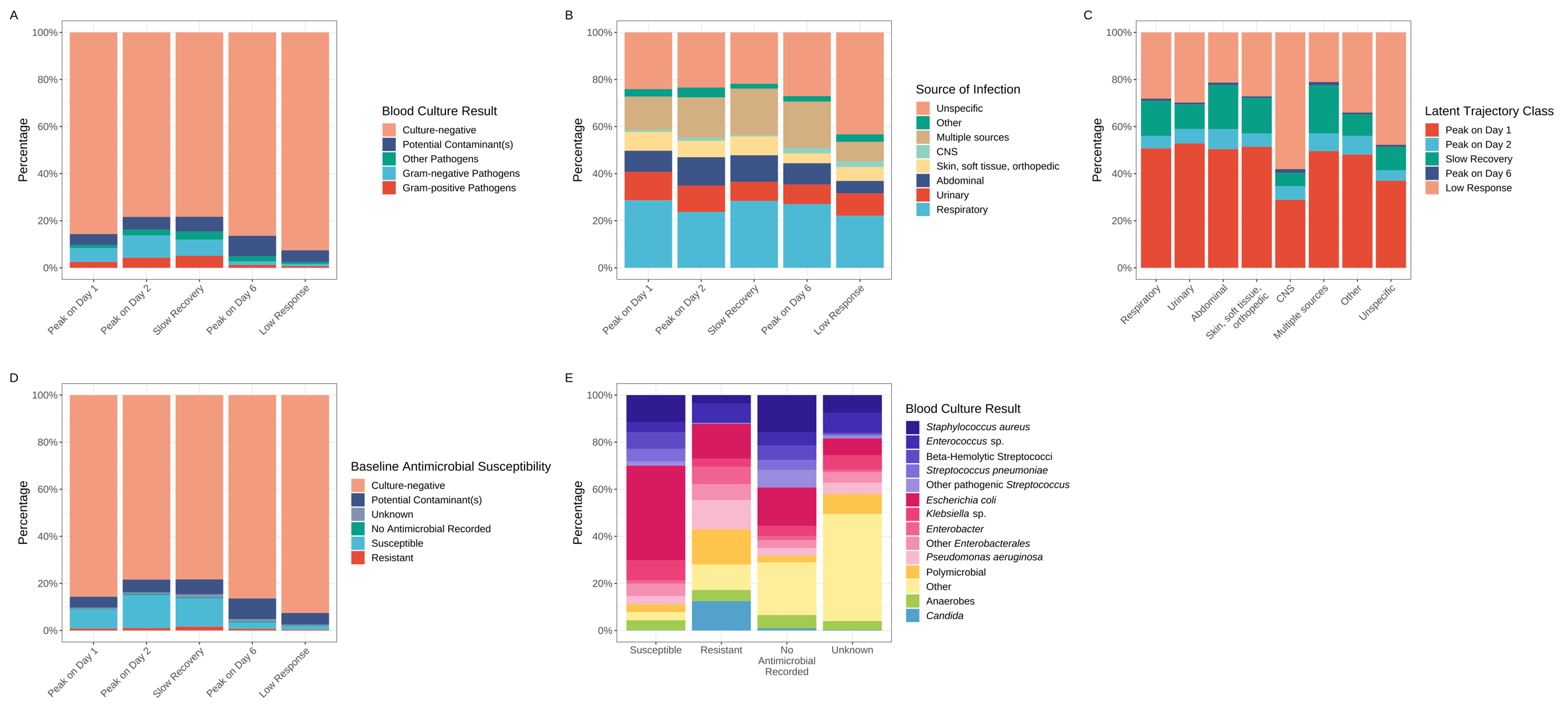

**Figure S9.** Associations between latent trajectory classes of the CRP response and blood culture results (A), sources of infection (B, C), baseline antimicrobial susceptibility (D), and pathogen groups' association with baseline antimicrobial susceptibility (E). See **Table S2** for cross table of pathogen groups versus baseline antimicrobial susceptibility.

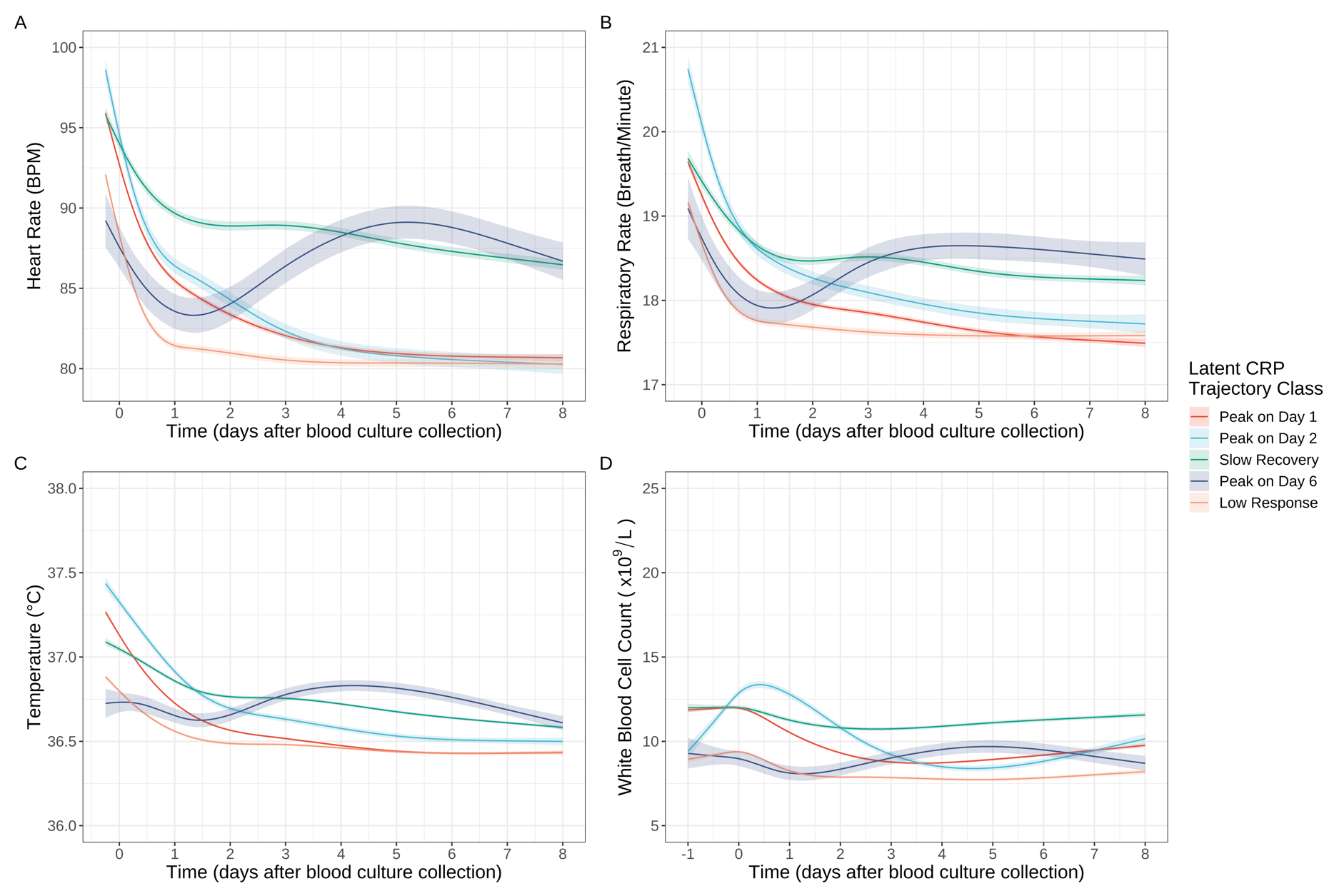

**Figure S10.** Response trajectories for heart rate (A), respiratory rate (B), temperature (C) and WBC count (D), by latent CRP trajectory class

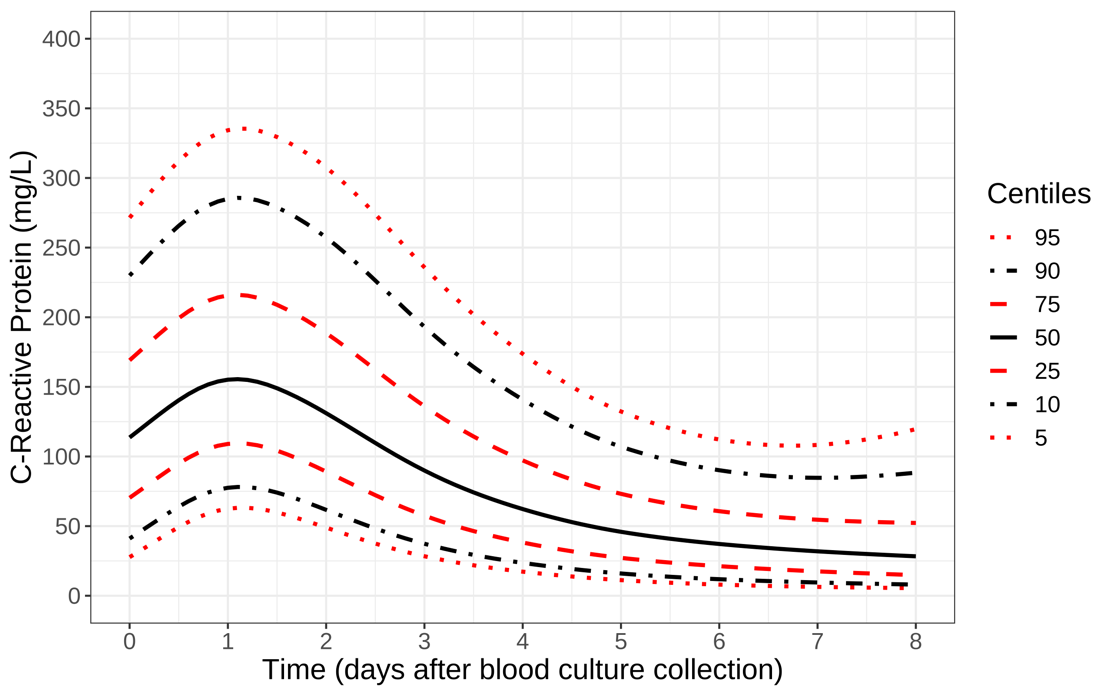

**Figure S11.** Centile reference chart estimated by selecting one random observation for each episode from those peaking on day 1 and day 2, regardless of pathogen isolated.

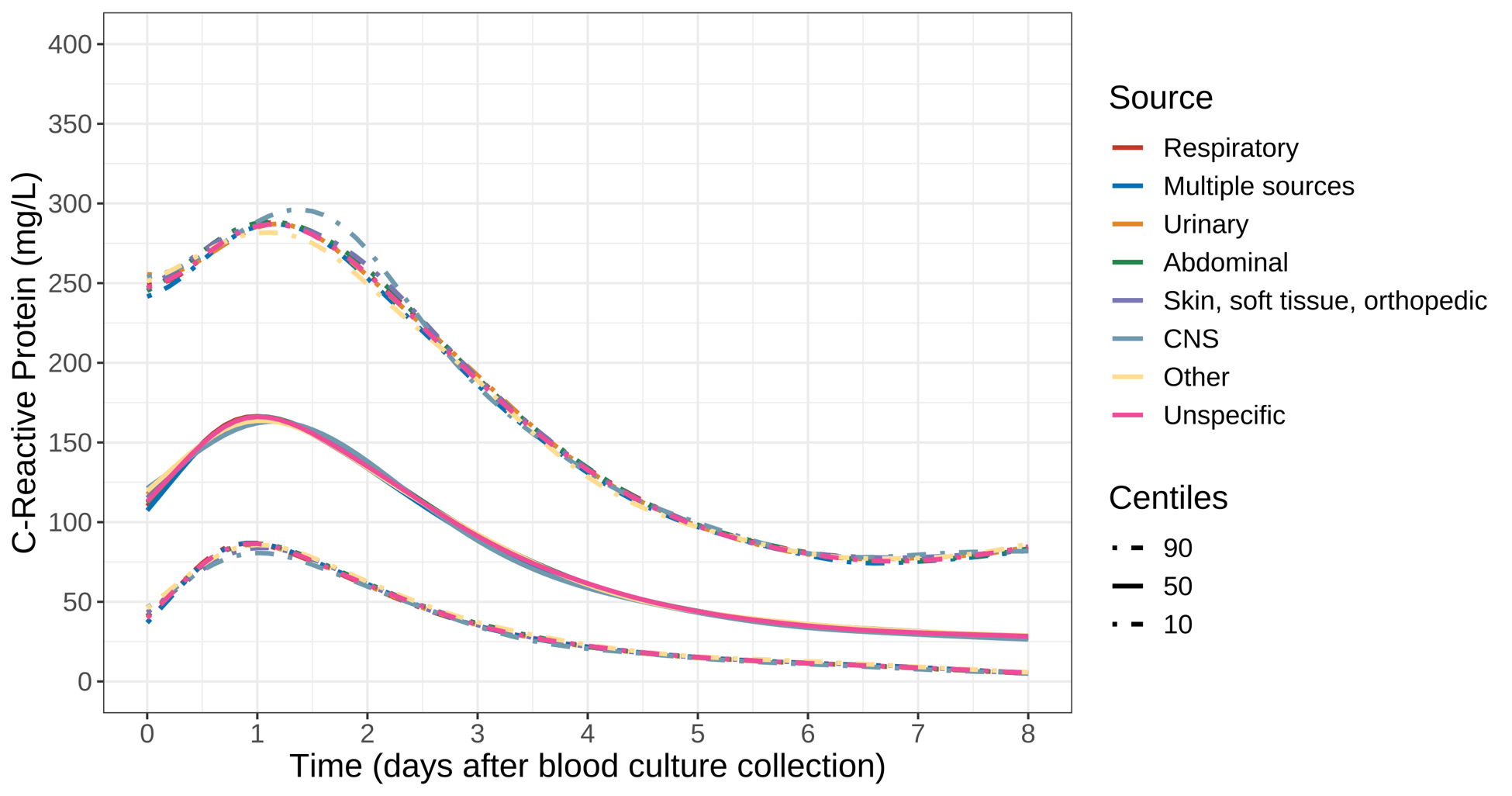

**Figure S12.** Centiles of expected CRP response in patients with culture-positive/negative suspected BSI following different sources of infection. Note: from the two latent classes peaking on day 1 and 2 in **Figure 2**, centiles for different sources of infection were estimated separately based on relevant episode subgroups, regardless of pathogen isolated.
